# Comorbidities and disability trajectories in multiple sclerosis: A two-cohort study using multi-state Markov models

**DOI:** 10.64898/2026.05.29.26354451

**Authors:** Chen Hu, Wen Zhu, Alexis Watterson, Sara Morini, Michele Morris, Shyam Visweswaran, Joyce Chang, Tianxi Cai, Tanuja Chitnis, Zongqi Xia

## Abstract

**Background:** Comorbidities are common in multiple sclerosis (MS) and may influence disability outcomes, but their dynamic impact on bidirectional disability transitions and long-term disability remains incompletely understood. Better understanding of this longitudinal relationship could inform personalized disability management strategies for people with MS.

**Methods:** We leveraged two large electronic health record (EHR)-linked MS registries and applied multi-state Markov models (MSMs) to examine the extent to which individual comorbidities and overall comorbidity burden were associated with short-term disability transitions, long-term disability transition probabilities, and expected time spent in each disability state. We additionally compared MSM-based predictions of confirmed disability worsening (CDW) with Cox proportional hazards (CoxPH) model-based predictions using the integrated Brier score with bootstrap validation.

**Results:** Among 3,723 patients with MS (74.6% female; 86.2% non-Hispanic White; mean age=41.9 years; mean disease duration=5.4 years) contributing 41,860 disability assessments over a mean follow-up of 7.3 years, higher cardiometabolic and psychiatric comorbidity burden was associated with increased transition intensity toward worse disability states and decreased transition intensity toward improvement, with a stepwise gradient across burden levels. Compared with patients without comorbidities, those with ≥4 comorbidities had a 28% higher risk of worsening (HR=1.28 [1.06, 1.55]) and a 20% lower risk of improvement (HR=0.80 [0.67, 0.95]). Each individual comorbidity was significantly associated with worse disability transitions. Long-term estimates indicated a higher 5-year probability of severe disability and fewer years spent in the no-disability state among patients with greater comorbidity burden. CoxPH models showed directionally consistent associations but lower predictive accuracy for CDW compared with MSMs.

**Conclusion:** Cardiometabolic and psychiatric comorbidities are associated with worse disability trajectories in MS, reducing improvement and accelerating progression. By providing a nuanced framework to quantify short-term disability transitions and long-term disability patterns, MSMs may have real-world clinical utility in disability prediction.

## Introduction

Multiple sclerosis (MS) is a chronic neuroimmune disease characterized by considerable variability in disease-related disability across individuals over time.^1–4^ Disability in MS is most commonly quantified using the Expanded Disability Status Scale (EDSS), a clinician-rated outcome, and the Patient-Determined Disease Steps (PDDS), a patient-reported outcome.^5–7^ While disease-specific factors such as age at symptom onset, relapse frequency, clinical subtype, and disease-modifying therapies (DMTs) explain some of this heterogeneity, patient-specific characteristics, including genetics, socioeconomic status, and comorbidities, are important additional contributors.^8^ Specifically, comorbidities are more modifiable through prevention or treatment than genetic or socioeconomic factors, offering actionable targets to improve MS management and prognosis.

Prior observational studies identified comorbidities, particularly psychiatric and cardiometabolic conditions, as associated with worse disability outcomes in MS.^9–16^ A meta-analysis of 17 clinical trials reported that higher comorbidity burden was linked to accelerated disability progression.^17^ However, the existing literature has notable gaps. First, prior investigations largely focused unidirectionally on disability worsening, overlooking the bidirectional (worsening and improving) and non-linear nature of disability dynamics in people with MS.^18–21^ A better understanding of how comorbidities influence bidirectional transitions could inform more personalized clinical management. Second, the exact timing of disability transitions is often unobserved, since disability assessments typically occur at discrete visits. Conventional survival analysis methods such as Cox proportional hazard (Cox PH) models are insufficient to address this scenario.^22^ Third, long-term disability trajectories across different levels of comorbidity burden are poorly understood. Cox PH models focus on a single time-to-event endpoint and neither accommodate intermediate states nor quantify the duration spent within each state, limiting their utility for examining longitudinal disability trajectories. Multi-state Markov models (MSMs) provide a flexible framework to model transitions between clinically defined discrete disability states over time, explicitly accounting for bidirectional change, intermittent observation, and time-varying covariates. MSMs have been applied in MS research to estimate transition rates, characterize the natural history of disease, compare disability progression patterns across phenotypes, and evaluate treatment effectiveness.^18,19,23–25^ However, relatively few studies have used this framework to examine how comorbidities influence disability dynamics beyond traditional single-endpoint analyses.

Methodologically, MSMs enable derivation of time-to-event probabilities, including survival curves for common endpoints such as 3-month confirmed disability worsening (CDW).^26,27^ This is feasible through their underlying transition probability structure. Operationally, CDW predictions from MSMs can be compared with those from Cox PH models using the same predictors.^28^ However, Cox PH models and MSMs are designed for different purposes. Cox models estimate the hazard of a single event, whereas MSMs model the full multi-state transition process simultaneously. Consequently, CDW probabilities derived from MSMs incorporate the broader structure of disability dynamics, whereas Cox models do not. Thus, comparing CDW predictions across the two frameworks should not be interpreted as a head-to-head test of model quality, but rather as an illustration of the additional information gained when prediction is embedded within a model structure that more closely reflects the underlying disease process. Whereas transition intensity ratios estimated from MSMs are conceptually analogous to hazard ratios from Cox PH models, MSMs enable simultaneous estimation of all possible transitions across states within a unified framework rather than modeling a single endpoint. Importantly, long-term dynamic measures cannot be directly derived from standard Cox PH models, which do not estimate the full transition probability structure across multiple states.

In this cohort study leveraging two electronic health record (EHR)-linked registries, we assessed short-term and long-term associations between selected comorbidities (three psychiatric and seven cardiometabolic conditions) and MS disability course using MSMs. By integrating registry and EHR data, comorbidity and covariate ascertainment is more reliable than with self-report, and the resulting real-world evidence extends beyond clinical trial findings. First, we tested whether comorbidities were associated with higher short-term transition intensities toward worse disability states and lower transition intensities toward improved states. Second, we quantified long-term differences across comorbidity burden groups, including 5-year state occupation probabilities and expected time spent in each disability state. Finally, we compared CDW predictions derived from MSMs with those from Cox PH models to assess how embedding prediction within a multi-state framework can yield estimates that more closely align with observed outcomes.

## Methods

### Data sources

This retrospective cohort study used two clinic-based MS registries, each linked with its corresponding EHR data (MGB EHR data from January 1, 1994, to December 31, 2022; UPMC EHR data from January 1, 2004, to December 31, 2024). Both EHR datasets contained structured data, including diagnosis and procedure codes (using the International Classification of Diseases [ICD] and Current Procedural Terminology [CPT] systems), healthcare utilization metrics (e.g., total counts of ICD codes and encounter notes), and medication history (e.g., RxNorm electronic prescriptions). We mapped all ICD codes to PheCodes using a published disease classification system to capture clinically meaningful phenotypes,^29^ consolidated CPT codes using the Clinical Classifications Software (CCS) for Services and Procedures, and grouped electronic prescription records at the RxNorm ingredient level.

We obtained EHR-linked registry data from the Prospective Investigation of Multiple Sclerosis in the Three Rivers Region (PROMOTE: Pittsburgh, PA) between 2017 and 2025, and the Comprehensive Longitudinal Investigation of Multiple Sclerosis at Brigham and Women’s Hospital (CLIMB: Boston, MA) between 2003 and 2025.^30,31^ Registries provided demographic information (e.g., age, sex, race, and ethnicity) and routinely collected clinical information (e.g., symptom onset, MS diagnosis date, relapse history, MS phenotypes, disability assessments, and DMTs). The primary disability outcome measure was the clinician-rated Expanded Disability Status Scale (EDSS) in CLIMB, and the patient-reported Patient Determined Disease Steps (PDDS) in PROMOTE.

### Ethics approval

The institutional review boards approved the study (University of Pittsburgh: STUDY19080007; Mass General Brigham: 1999P010435). All registry participants provided written informed consent prior to enrollment.

### Study population

Eligibility criteria included (1) neurologist-confirmed MS diagnosis; (2) age ≥18 years at MS symptom onset; (3) ≥3 years of continuous care within the UPMC or MGB healthcare systems, defined as no gap longer than one year between any two EHR entries; (4) first neurologist-confirmed MS diagnosis recorded during the observation window, defined as the interval between the first and last EHR entry with evidence of continuous care; and (5) ≥2 disability assessments (EDSS for CLIMB and PDDS for PROMOTE) during the observation window, with a minimum interval of three months between assessments.

From both cohorts, we extracted data on date of birth, sex, race and ethnicity, healthcare utilization, dates of symptom onset and MS diagnosis, MS phenotype, relapse counts, DMT details (i.e., drug names and treatment start and end dates), and all recorded disability scores with corresponding assessment dates. Participants with missing data for any of these key variables were excluded from the analysis. A high-level overview of the study design and key concepts is presented in **Figure 1A**. A summary of the cohort construction process is shown in **Figure 1B**. The final analysis included 1,008 individuals from PROMOTE and 2,715 individuals from CLIMB, contributing a total of 9,294 and 32,566 qualified PDDS and EDSS assessments, with mean follow-up durations of 4.2 years and 8.4 years, respectively.

**Figure 1.**
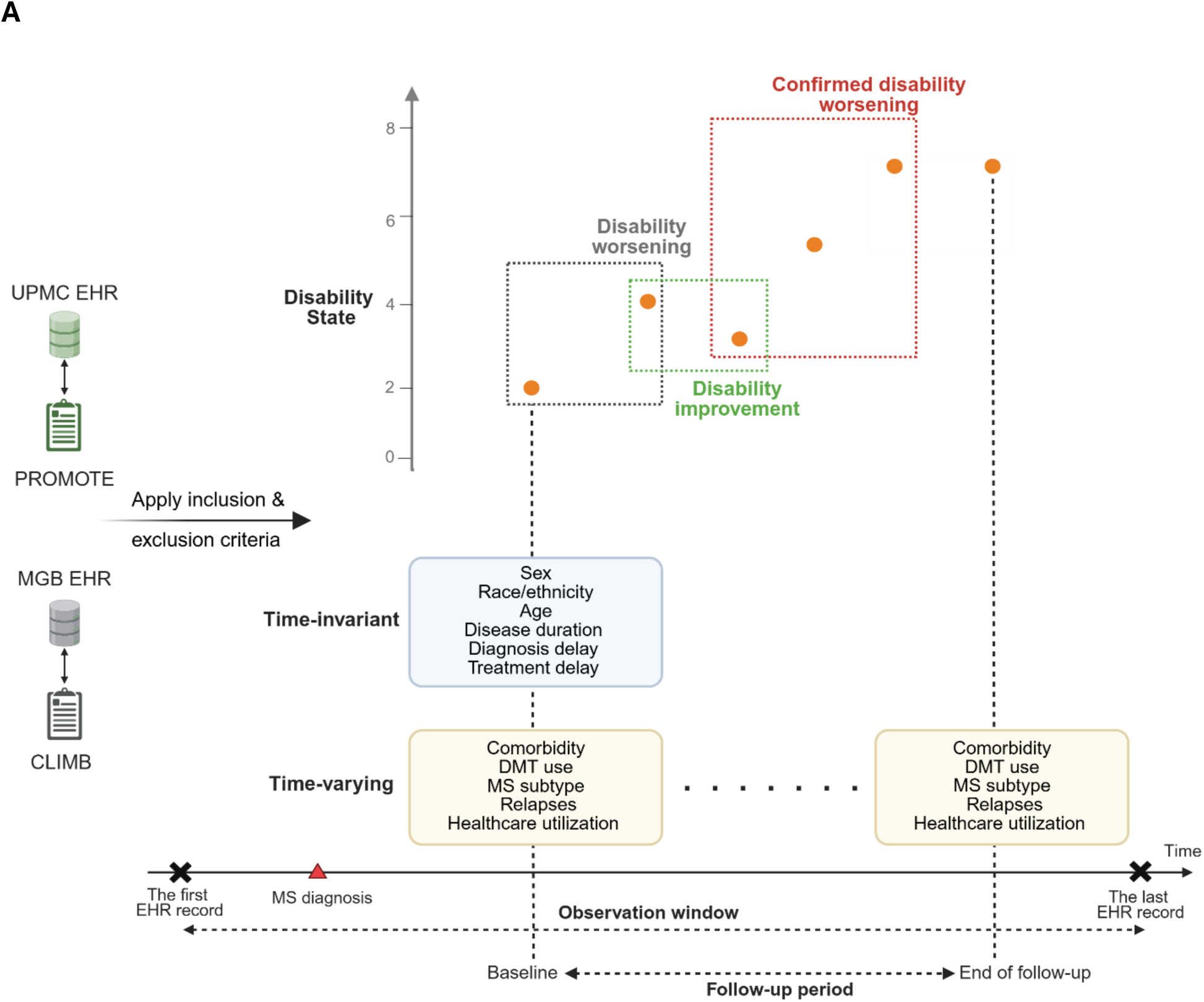
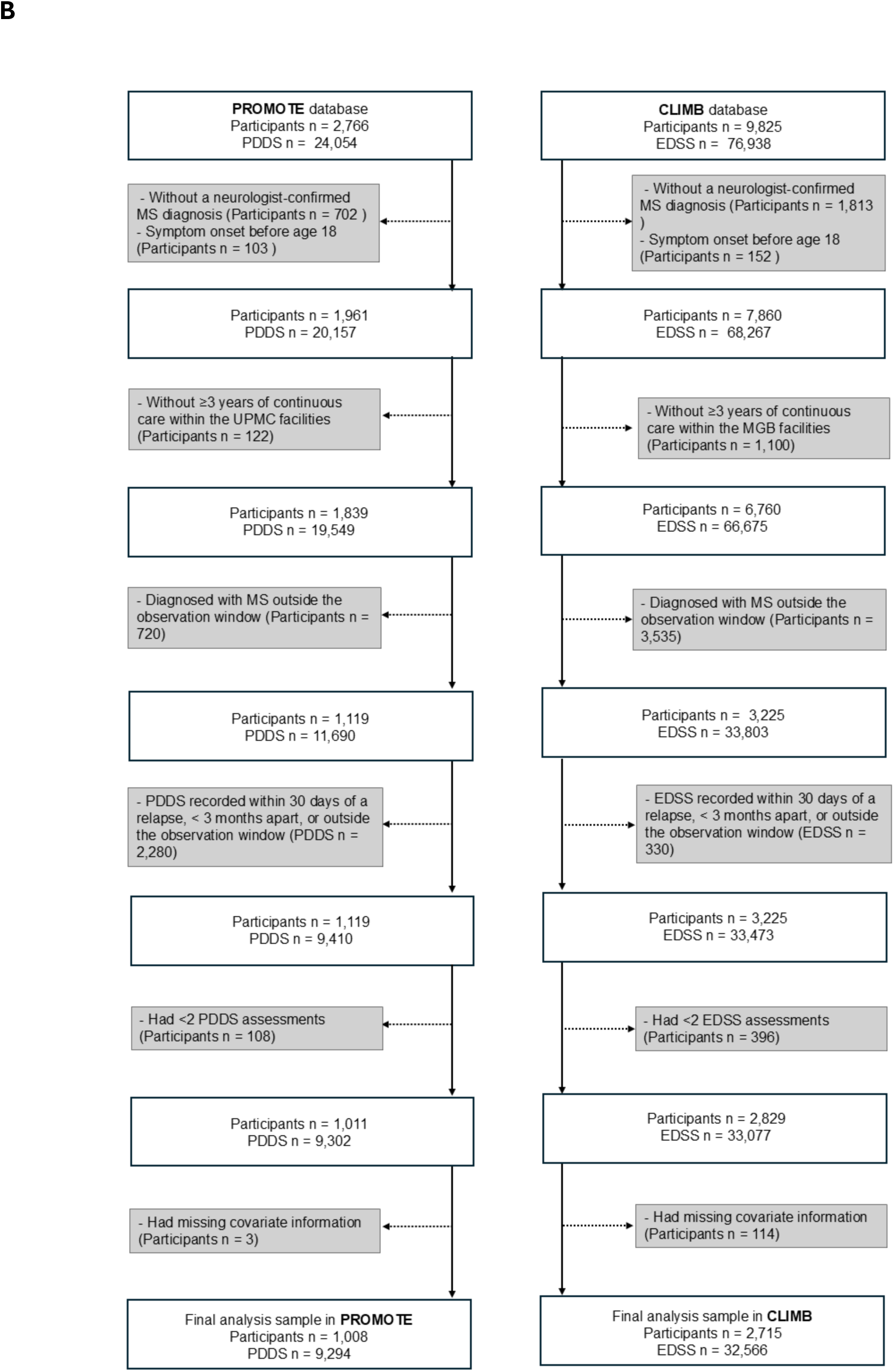
Study design. (A) Schematic overview. (B) Flowchart of participant selection. MS, multiple sclerosis; EHR, electronic health record; PROMOTE, the Prospective Investigation of Multiple Sclerosis in the Three Rivers Region; CLIMB, the Comprehensive Longitudinal Investigation of Multiple Sclerosis at Brigham; PDDS, patient determined disease Steps; EDSS, expanded disability status scale.

### Comorbidities

#### Individual comorbidities

We examined the presence of chronic cardiometabolic and psychiatric comorbidities recommended by the International Advisory Committee on Clinical Trials in MS.^8^ Cardiometabolic conditions included hypertension, hyperlipidemia, diabetes, cerebrovascular disease, functional cardiovascular disorders, ischemic heart disease, and peripheral vascular disease. Psychiatric conditions included depression, anxiety, and other mental health disorders (e.g., bipolar disorder, personality disorder, and alcohol, tobacco, or substance use disorders). We identified comorbidities within each participant’s observation window using PheCodes (**eTable 1**). We mapped related ICD codes to PheCodes using a published system in which each PheCode represents a clinically distinct medical condition.^32^ A comorbidity was considered present if a participant had ≥2 occurrences of the corresponding PheCode, separated by at least three days. We treated comorbidity status as a time-varying variable. Once a participant met the criteria for a specific comorbidity, they were considered to have that condition for the remainder of the follow-up period, reflecting the chronic and often recurrent nature of these conditions. We excluded medication data from our comorbidity definitions to avoid misclassification that could result from off-label medication use.

#### Comorbidity burden

We defined the total comorbidity burden as the total number of comorbidities (maximum: 10) and categorized it as 0, 1, 2, 3, or ≥4. We also separately evaluated the burden of cardiometabolic conditions (hypertension, hyperlipidemia, diabetes, cerebrovascular disease, functional cardiovascular disease, ischemic heart disease, and peripheral vascular disease) and psychiatric conditions (depression, anxiety, and other psychiatric disorders), each categorized as 0, 1, or ≥2. We treated all comorbidity burden measures as time-varying variables.

### Disability outcomes

#### Disability states

The PDDS is an ordinal scale with nine levels ranging from 0 (normal) to 8 (bedridden). The EDSS ranges from 0 (normal neurological examination) to 10 (death due to MS) in 0.5 increments. Higher scores on both scales indicate greater disability. To reduce model complexity and limit the number of parameters, EDSS scores were rounded up to the nearest integer (e.g., a score of 3.5 was rounded to 4), while PDDS scores were used as originally recorded. Based on these processed scores, we categorized disability into nine states (State 0 through State 8), as described in **eTable 2**.

#### Disability worsening and improvement

Disability worsening was defined as an increase in the pre-defined disability state from one visit to the next, while disability improvement was defined as a decrease in the disability state (**Figure 1A**). We applied MSM models to simultaneously capture transitions toward both higher (worsening) and lower (improving) disability states (details in Statistical Analyses).

#### Confirmed disability worsening (CDW)

CDW is a widely used clinical endpoint in MS research, representing a sustained increase in disability that persists over time. In this study, 3-month CDW was defined as disability worsening relative to baseline that remained at a subsequent visit ≥3 months later (**Figure 1A**). This outcome has traditionally been analyzed using Cox PH models. In the present study, we used 3-month CDW to compare the predictive performance of MSM models versus Cox PH models.

### Covariates

We selected covariates *a priori* based on clinical judgment and prior literature.^25,33^ Time-invariant covariates were measured at baseline, defined as the date of the first available disability state assessment. Time-invariant covariates included sex (male vs. female), race/ethnicity (non-Hispanic White vs. other minorities), age, disease duration (years from symptom onset to baseline), diagnosis delay (years from symptom onset to MS diagnosis), and treatment delay (years from symptom onset to the earlier of either DMT initiation or baseline date). Time-varying covariates included DMT use, MS subtype (progressive MS vs. relapsing-remitting MS), total number of relapses, and healthcare utilization (log-transformation total number of ICD codes and clinical encounter notes). We defined DMT use as a categorical variable with three groups: no DMT, standard-efficacy DMTs, and higher-efficacy DMTs (**eMethod-1**).

### Statistical analyses

#### Descriptive analysis

We summarized participant characteristics using mean (SD), median (IQR), or frequency (proportion), as appropriate for each cohort. We reported pooled estimates as weighted means for continuous variables and weighted proportions for categorical variables.

#### Multi-state Markov model (MSM)

To assess the associations between comorbidities (individual condition or total burden) and transitions across disability states, we applied nine-state inhomogeneous, continuous-time MSM models, adjusting for all previously described covariates. Model terms are detailed in **eTable 3**. This approach assumes that transition rates between states vary over time (inhomogeneous) and can occur at any point (continuous time), a structure previously shown to be appropriate for modeling disability dynamics in MS.^23,25^ We modeled both disability worsening (transitions to higher states) and improvement (transitions to lower states) simultaneously, assuming that each participant occupies only one state at any given time. Following previous studies, direct transitions between non-adjacent states were not permitted, and individuals were required to pass sequentially through intermediate states.^23^ To balance model complexity with sample size and ensure convergence, we specified a progression-regression (PR) structure, assuming a common transition intensity for all progression transitions (e.g., state 0→1 through 8→9) and a common intensity for all regression transitions (e.g., state 1→0 through 9→8). We measured time to transition from the baseline disability state, defined as the first recorded disability assessment, aligning the multi-state process with the start of clinical follow-up. Participants were censored at their last available disability assessment. The use of clinic registry data minimized the likelihood of informative censoring.

This structure constrains comorbidity effects to be homogeneous across all state transitions, which may not fully reflect clinical reality, as comorbidities could differentially influence early-versus late-stage disability transitions. Accordingly, we assessed model fit under these pre-specified assumptions using both graphical diagnostics and a Pearson-type goodness-of-fit test. In the diagnostic plots, we compared observed and model-predicted proportions of patients occupying each disability state over time. The Pearson goodness-of-fit test provided a formal comparison between observed and expected counts across states. We considered model fit adequate when predicted values closely aligned with observed values in the plots and the goodness-of-fit test was not statistically significant (as the null hypothesis is that the models fit the data well).

To quantify associations between comorbidities and disability transitions, we reported transition intensity ratios and 95% credible intervals (CIs) for individual comorbidity status and composite comorbidity burden, separately for each cohort and then pooled across cohorts. These ratios represent the relative intensity of transitioning between states, conditional on covariate values, and are formally transition intensity ratios within the MSM framework rather than hazard ratios in the Cox PH sense. For consistency with reporting conventions in the MS literature and to facilitate comparison with Cox PH results, we refer to both transition intensity ratios from MSMs and hazard ratios from Cox PH models as hazard ratios (HRs) throughout. To account for estimation variability, we computed empirical 95% CIs by penalizing the standard errors of regression coefficients, following the approach described by Blume *et al.* (see **eMethod-2**).^34^ We obtained pooled estimates using fixed-effect meta-analysis and quantified between-cohort heterogeneity using the I² statistic, with values of 0–50% indicating low heterogeneity and 51–100% indicating high heterogeneity.^35^ Given that only two cohorts were available, we did not use random-effects models, as they would yield unstable estimates of between-study variance. For comorbidities with high between-cohort heterogeneity (I² > 50%), pooled estimates should be interpreted with caution, as the fixed-effects assumption of a single underlying true effect may not hold. In these cases, cohort-specific estimates are more informative.

To estimate long-term disability trajectories, we calculated the probability of entering each state over a 5-year follow-up, and the mean state occupation times at the end of the 5-year follow-up across five comorbidity burden levels (0, 1, 2, 3, and ≥4 comorbidities). Estimates were generated for a reference patient profile: a 40-year-old, non-Hispanic White female with newly diagnosed, treatment-naïve RRMS and one prior relapse. We selected this profile to reflect average cohort characteristics, the majority demographic composition, and a clinically meaningful scenario for interpretation.

#### Cox proportional hazard model (Cox PH)

In parallel, we used Cox PH models to examine associations between comorbidities (individual or total burden) and the risk of 3-month CDW. These models adjusted for the same covariates included in the MSM, with the addition of baseline disability state. This difference reflects the fact that MSMs inherently account for baseline state when modeling transition probabilities, whereas Cox PH models require explicit adjustment for it. We incorporated time-varying covariates using a start-stop (interval) data structure, in which each subject contributed multiple observation intervals that representing periods during which covariate values remained constant. Model terms are detailed in **eTable 4**. Patients who met the criteria for disability worsening but lacked a subsequent disability assessment were censored at their last visit before meeting the worsening criteria. We report HRs and 95% CIs for individual comorbidity status and composite comorbidity burden. HRs represent the relative risk of 3-month CDW, conditional on covariate values.

#### Model performance in predicting 3-month confirmed disability worsening

To illustrate the practical implications of model choice, we examined the predicted cumulative risk of 3-month CDW derived from both the MSM and Cox PH models. Because these models differ fundamentally in structure and purpose, this comparison is *not* intended as a head-to-head evaluation of competing approaches, but rather as a demonstration of how embedding CDW prediction within a richer transition framework may yield predictions that more closely align with observed outcomes. Both models included all individual comorbidities, demographic variables, and clinical factors as predictors. We assessed model performance using the integrated Brier score (**IBS**), which represents the summary weighted squared difference between predicted and observed survival probabilities.^36^ Inverse probability censoring weighting was used to adjust for censoring. We obtained 95% CIs for the IBS via nonparametric bootstrap with 500 resamples. To compute MSM-based predictions of 3-month CDW, we first used the MSM-estimated transition intensities to obtain patient-specific transition probabilities given their current covariate values. We then applied the method described in Mandel *et al.* (**eMethod-3**) to convert these transition probabilities into predicted probabilities of 3-month CDW.^37^ For the Cox PH model, we obtained predicted probabilities of 3-month CDW directly from the fitted model by providing baseline values for time-constant predictors and observed trajectories for time-varying predictors.

#### Sensitivity analyses

Because of limited pre-baseline EHR data, we anticipated potential misclassification: participants with pre-existing comorbidities might be incorrectly classified as not having them. To assess whether this misclassification could bias our estimates, we conducted a sensitivity analysis in which 5% and 20% of participants without each comorbidity at baseline were randomly reassigned to the presence group, and MSM models were refitted.^38^

#### Software

All statistical analyses were performed with R (version 4.4.1). We used the *msm* package to fit the MSM model.^39^ We provide the codes for analyses and figures: https://github.com/xialab2016/Aim-2-Comorbidities-and-disability-transitions.

### Data sharing statement

Sharing of de-identified EHR and registry data with qualified external researchers may be permissible with approval from the IRB and regulatory oversight agents of the healthcare system once an appropriate Data Use Agreements (DUA) between institutions is in place following reasonable request to the corresponding author.

## Results

### Descriptive statistics

3,723 patients (PROMOTE: n=1,008, CLIMB: n=2,715; 74.6% female; 86.2% non-Hispanic White; mean age at baseline 41.9 years; mean disease duration at baseline 5.4 years; 71.2% in less advanced disability state of 0–2) met the inclusion criteria, contributing 41,860 disability assessments over a mean follow-up of 7.3 years (**Table 1**). The pooled prevalence of individual comorbidities was largely consistent with prior reports in non–clinical trial populations: anxiety (23.9%) and depression (22.7%) were the most common, followed by hypertension (18.2%) and hyperlipidemia (16.3%).

**Table 1.** Demographics and clinical characteristics of participants.

|  | PROMOTE | CLIMB | Pooled statistics <sup>1</sup><br>(95%CI) |
| --- | --- | --- | --- |
| Number of participants | 1,008 | 2,715 | 3,723 |
| Number of disability assessments | 9,294 | 32,566 | 41,860 |
| **Baseline characteristics** |  |  |  |
| Female; n (%) | 770 (76.4%) | 2,006 (73.9%) | 74.6 (73.1, 75.9) |
| Non-Hispanic White; n (%) | 879 (87.2%) | 2,341 (86.2%) | 86.2 (85.1, 87.3) |
| Age (Years); Mean (SD) | 44.7 (12.3) | 40.8 (11.6) | 41.9 (41.0, 42.7) |
| MS duration (Years); Mean (SD) | 8.6 (8.1) | 4.2 (6.3) | 5.4 (4.9, 5.9) |
| Diagnosis delay (Years); Mean (SD) | 3.7 (6.0) | 3.5 (5.8) | 3.6 (3.1, 4.0) |
| Progressive MS; n (%) | 89 (8.8%) | 237 (8.7%) | 8.8 (7.9, 9.7) |
| DMT use; n (%) |  |  |  |
| Never | 258 (25.6%) | 682 (25.1%) | 25.2 (23.9, 26.7) |
| Ever used standard-efficacy DMTs | 605 (60.0%) | 1,824 (67.2%) | 65.2 (63.7, 66.8) |
| Ever used high-efficacy DMTs | 343 (34.0%) | 343 (12.6%) | 18.4 (17.2, 19.7) |
| Treatment delay (Years); Mean (SD) | 4.5 (6.6) | 3.5 (5.9) | 3.8 (3.3, 4.2) |
| Number of prior relapses; Mean (SD) | 1.1 (1.7) | 1.4 (0.9) | 1.3 (1.2, 1.4) |
| Healthcare utilization; Mean (SD) | 7.8 (0.7) | 9.3 (0.9) | 8.9 (8.8, 9.0) |
| Disability state; n (%) |  |  |  |
| State 0 | 384 (38.1%) | 584 (21.5%) | 26.0 (24.6, 27.4) |
| State 1 | 219 (21.7%) | 506 (18.6%) | 19.5 (18.2, 20.8) |
| State 2 | 119 (11.8%) | 838 (30.9%) | 25.7 (24.3, 27.1) |
| State 3 | 96 (9.5%) | 364 (13.4%) | 12.4 (11.3, 13.5) |
| State 4 | 83 (8.2%) | 196 (7.2%) | 7.5 (6.7, 8.4) |
| State 5 | 62 (6.2%) | 29 (1.1%) | 2.4 (2.0, 3.0) |
| State 6 | 22 (2.2%) | 106 (3.9%) | 3.4 (2.9, 4.1) |
| State 7 | 22 (2.2%) | 63 (2.3%) | 2.3 (1.9, 2.8) |
| State 8 | 1 (0.1%) | 29 (1.1%) | 0.8 (0.6, 1.1) |
| Total comorbidity burden; n (%) |  |  |  |
| 0 | 448 (44.4%) | 1,533 (56.5%) | 53.2 (51.6, 54.8) |
| 1 | 210 (20.8%) | 474 (17.5%) | 18.4 (17.1, 19.6) |
| 2 | 142 (14.1%) | 305 (11.2%) | 12.0 (11.0, 13.1) |
| 3 | 104 (10.3%) | 189 (7.0%) | 7.9 (7.0, 8.8) |
| ≥4 | 104 (10.3%) | 214 (7.9%) | 8.5 (7.7, 9.5) |
| Cardiometabolic burden; n (%) |  |  |  |
| 0 | 646 (64.1%) | 1,939 (71.4%) | 69.4 (67.9, 70.9) |
| 1 | 192 (19.0%) | 399 (14.7%) | 15.9 (14.7, 17.1) |
| ≥2 | 170 (16.9%) | 377 (13.9%) | 14.7 (13.6, 15.9) |
| Psychiatric burden; n (%) |  |  |  |
| 0 | 606 (60.2%) | 1,901 (70.0%) | 67.3 (65.8, 68.8) |
| 1 | 201 (19.9%) | 423 (15.6%) | 16.8 (15.6, 18.0) |
| ≥2 | 201 (19.9%) | 391 (14.4%) | 15.9 (14.8, 17.1) |
| Depression; n (%) | 276 (27.4%) | 570 (21.0%) | 22.7 (21.4, 24.1) |
| Anxiety; n (%) | 307 (30.5%) | 583 (21.5%) | 23.9 (22.6, 25.3) |
| Other psychiatric disorders; n (%) | 61 (6.1%) | 110 (4.1%) | 4.6 (4.0, 5.3) |
| Hypertension; n (%) | 223 (22.1%) | 454 (16.7%) | 18.2 (17.0, 19.5) |
| Hyperlipidemia; n (%) | 215 (21.3%) | 392 (14.4%) | 16.3 (15.2, 17.5) |
| Cerebrovascular disease; n (%) | 21 (2.1%) | 31 (1.1%) | 1.4 (1.1, 1.8) |
| Functional cardiovascular disease; n (%) | 73 (7.2%) | 188 (6.9%) | 7.0 (6.2, 7.9) |
| Ischemic heart disease; n (%) | 33 (3.3%) | 123 (4.5%) | 4.2 (3.6, 4.9) |
| Peripheral vascular disease; n (%) | 31 (3.1%) | 105 (3.9%) | 3.7 (3.1, 4.3) |
| Diabetes; n (%) | 90 (8.9%) | 153 (5.6%) | 6.5 (5.8, 7.4) |
| **Follow-up (FU) characteristics** |  |  |  |
| FU years; Mean (SD) | 4.2 (2.2) | 8.4 (5.9) | 7.3 (6.9, 7.6) |
| Observation window (Years); Mean (SD) | 19.7 (8.4) | 17.4 (5.9) | 18.0 (17.5, 18.5) |
| Total relapses during FU; Mean (SD) | 0.7 (0.8) | 1.6 (1.4) | 1.4 (1.3, 1.5) |
| Disability worsening during FU; n (%) | 738 (73.2%) | 1,942 (71.5%) | 0.72 (0.71, 0.73) |
| Disability improvement during FU; n (%) | 717 (71.1%) | 1,715 (63.2%) | 0.65 (0.64, 0.67) |
| 3-month CDW during FU; n (%) | 445 (44.1%) | 1,630 (60.0%) | 0.56 (0.54, 0.57) |
| 6-month CDW during FU; n (%) | 403 (40.0%) | 1,554 (57.2%) | 0.52 (0.50, 0.54) |

<sup>1</sup> Pooled statistics are weighted means for continuous variables and weighted proportions for categorical variables.
Abbreviation: PROMOTE, the Prospective Investigation of Multiple Sclerosis in the Three Rivers Region; CLIMB, the Comprehensive Longitudinal Investigation of Multiple Sclerosis in Brigham; DMT, disease-modifying therapy; CDW, confirmed disability worsening

Compared with CLIMB participants, PROMOTE participants were older (44.7 vs. 40.8 years), had longer disease duration (8.6 vs. 4.2 years), and had a higher proportion who ever used high-efficacy DMTs (34.0% vs. 12.6%). Otherwise, the two cohorts had comparable sex, race and ethnicity, diagnostic delay, baseline MS subtype, treatment delay, number of relapses before baseline, and health care utilization. Baseline comorbidity burden was generally greater in PROMOTE than in CLIMB: 44.4% of PROMOTE vs. 56.5% of CLIMB participants had no studied comorbidities; 64.1% vs. 71.4% had no cardiometabolic comorbidities; and 60.2% vs. 70.0% had no psychiatric comorbidities.

The distribution of disability states differed between cohorts, reflecting inherent differences between the PDDS (PROMOTE) and EDSS (CLIMB) measurements. During follow-up (PROMOTE: mean 4.2 years; CLIMB: mean 8.4 years), disability worsening occurred in 73.2% of PROMOTE participants and 71.5% of CLIMB participants, while disability improvement occurred in 71.1% and 63.2%, respectively. The proportion reaching the 3-month CDW milestone was lower in PROMOTE (44.1%) than in CLIMB (60.0%), largely due to shorter disability assessment follow-up. Consequently, this study focused on changes in disability over time rather than absolute disability at a single time point.

### Model assessment

As shown in the diagnostic plots (**eFigure 1A-B**), the base model (adjusted for all covariates except comorbidity variables) showed good fit in both cohorts, with acceptable agreement between observed and expected prevalence of state occupancies over time. The Pearson-type goodness-of-fit test further supported adequate model fit, yielding non-significant p-values of 0.45 in PROMOTE and 0.34 in CLIMB. We did not conduct additional goodness-of-fit assessments for models including each individual comorbidity or composite burden, as the satisfactory performance of the base model supported the pre-specified modeling structure and assumptions, including the nine-state framework, restriction to adjacent transitions, definition of time scale, and progression-regression assumption.

### Comorbidities and disability transitions using MSM models

The presence of comorbidities was associated with both greater risk of disability worsening (**Figure 2A, eTable 5**) and reduced likelihood of disability improvement (**Figure 2B, eTable 6**), after adjusting for sex, race/ethnicity, age, disease duration, diagnosis delay, treatment delay, healthcare utilization, DMT use, MS subtype, and total number of relapses. Compared with patients without any of the studied comorbidities, those with 3 and ≥4 comorbidities had a 16% higher hazard of disability worsening (HR = 1.16, 95% CI: 1.04-1.30) and a 28% higher hazard (HR = 1.28, 95% CI: 1.06-1.55), respectively, while experiencing a 5% (HR = 0.95, 95% CI: 0.92-0.99) and 20% (HR = 0.80, 95% CI: 0.67-0.95) lower likelihood of disability improvement. Having 1 or 2 comorbidities was also associated with disability progression (HR = 1.12, 95% CI: 1.03-1.22, and HR = 1.15, 95% CI: 1.03-1.27, respectively), though not with improvement.

**Figure 2.**
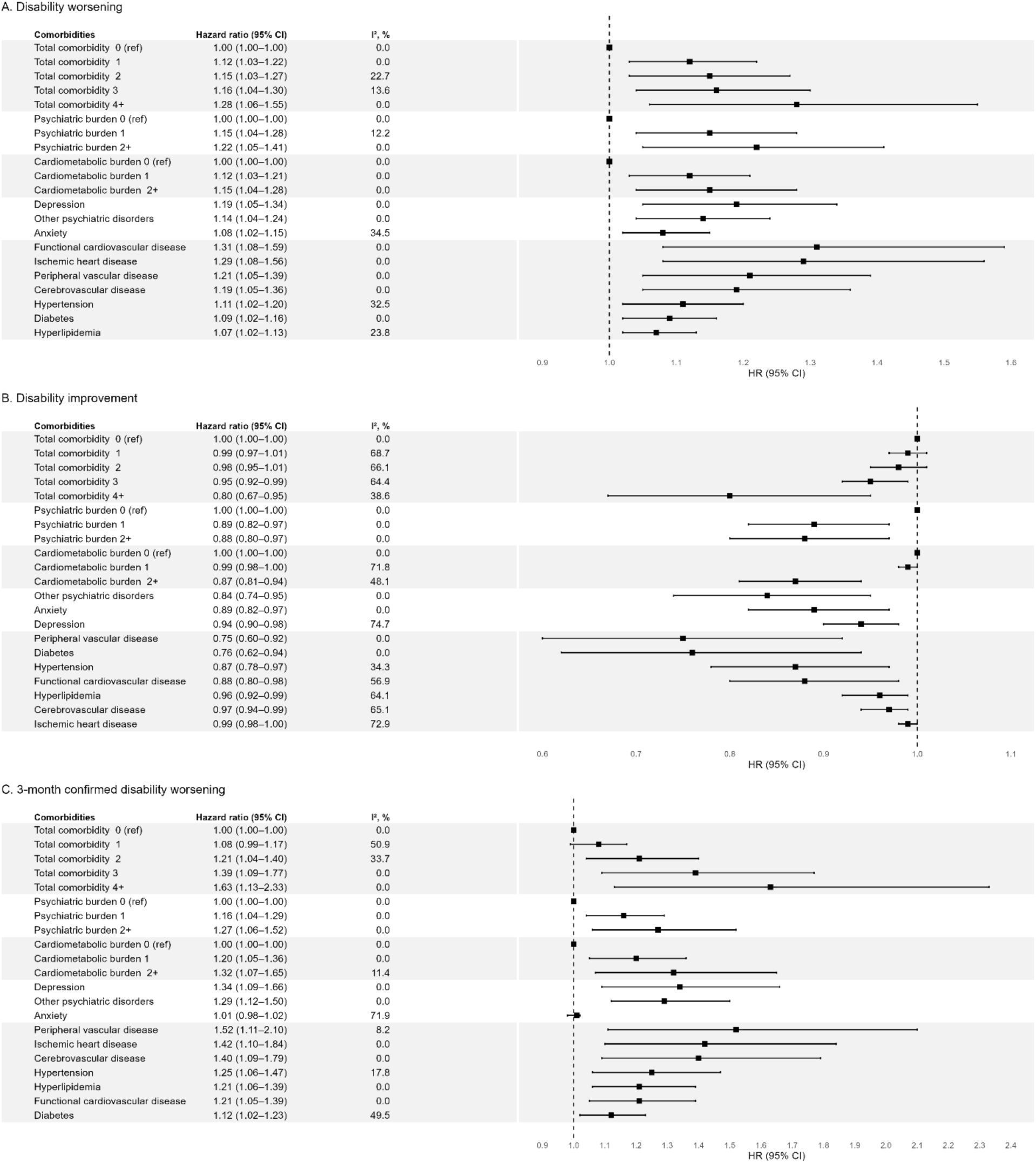
Pooled associations between comorbidities and disability outcomes: (A) Disability worsening (B) Disability improvement, and (C) 3-month confirmed disability worsening. All models adjusted for included comorbidities, sex, race/ethnicity, age, disease duration, diagnosis delay, treatment delay, healthcare utilization, DMT use, MS subtype, and total number of relapses.

Psychiatric comorbidity burden was consistently linked to adverse outcomes. Compared with patients without any psychiatric comorbidity, having 1 psychiatric disorder was associated with a 15% increased hazard of disability worsening (HR = 1.15, 95% CI: 1.04-1.28) and an 11% lower likelihood of improvement (HR = 0.89, 95% CI: 0.82-0.97). The presence of ≥2 psychiatric conditions was associated with even greater disability worsening (HR = 1.22, 95% CI: 1.05–1.41) and a lower likelihood of improvement (HR = 0.88, 95% CI: 0.80-0.97).

Cardiometabolic comorbidity burden showed similar associations. Having 1 cardiometabolic condition was associated with higher risk of worsening (HR = 1.12, 95% CI: 1.03–1.21) and borderline lower improvement (HR = 0.99, 95% CI: 0.98-1.00), while ≥2 conditions were associated with both increased worsening (HR = 1.15, 95% CI: 1.04–1.28) and reduced improvement (HR = 0.87, 95% CI: 0.81–0.94).

When examined individually, all comorbidities were significantly associated with disability outcomes. Hazard ratios for worsening ranged from 1.07 for hyperlipidemia to 1.31 for functional cardiovascular disease, indicating a higher likelihood of transitioning to a worse disability state; hazard ratios for improvement ranged from 0.75 for peripheral vascular disease to 0.99 for ischemic heart disease, suggesting a lower likelihood of transitioning to a better disability state.

Heterogeneity between cohorts was generally low for worsening transitions (I² range 0-34.5%) but was substantially higher for several improvement transitions, particularly for depression (I² = 74.7%), ischemic heart disease (I² = 72.9%), cardiometabolic burden level 1 (I² = 71.8%), and total comorbidity burden levels 1-3 (I² = 64.4–68.7%). For these estimates, pooled HRs should be interpreted with caution. Cohort-specific results in **eTables 5 and 6** are more informative.

### Long-term disability trajectories stratified by total burden of comorbidities using MSM models

For a hypothetical 40-year-old, non-Hispanic White, treatment-naïve female with newly diagnosed RRMS, one prior relapse, and no current disability (state 0), the probability of reaching the most severe disability state (state 8) within 5 years was 6% among those without comorbidity and 13% among those with ≥4 comorbidities (**Figure 3**). In contrast, the probability of remaining in the no-disability state at 5 years was lower among individuals with ≥4 comorbidities than those without any comorbidity (15% vs. 29%). Overall, an increasing comorbidity burden was associated with a higher probability of transitioning to disability states above state 2 and a lower probability of remaining in the no-disability state (**eTable 7**). Expected time spent in each state over the 5-year follow-up is presented in **eTable 8**. Participants with ≥4 comorbidities spent fewer years in the healthy state (2.0 vs. 3.0 years) and more years in disability states at or above state 2 (2.2 vs. 1.5 years) compared with those without comorbidities.

**Figure 3.**
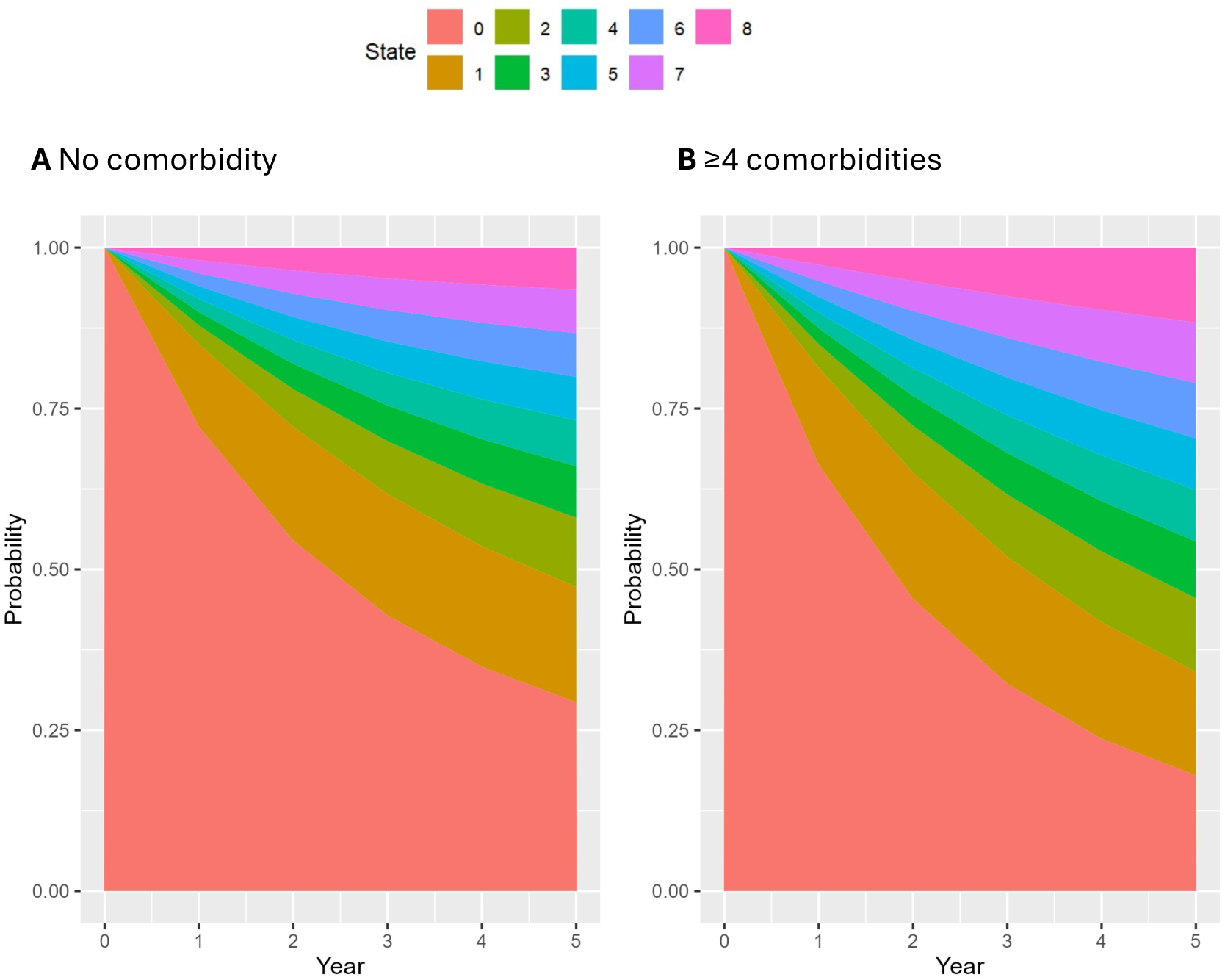
Stacked predicted state occupancy probabilities with time by comorbidity burden: (A) No comorbidity; (B) ≥4 comorbidities. Probabilities were predicted based on the Multi-state Markov (MSM) model for a hypothetical 40-year-old, non-Hispanic White, treatment-naïve female with newly diagnosed RRMS, one prior relapse, and no disability (state 0) at Year 0.

### Comorbidities and 3-month confirmed disability worsening (CDW) using Cox PH models

Using 3-month CDW (**Figure 2C, eTable 9**) as the endpoint and adjusting for covariates, patients with 2, 3, and ≥4 comorbidities had 21% (HR = 1.21, 95% CI: 1.04–1.40), 39% (HR = 1.39, 95% CI: 1.09–1.77), and 63% (HR = 1.63, 95% CI: 1.13–2.33) higher hazards of 3-month CDW, respectively, compared with those without comorbidities. Having one comorbidity was not significantly associated with CDW risk (HR = 1.08, 95% CI: 0.99–1.17). Psychiatric conditions were strongly associated with CDW risk. Having one psychiatric disorder was linked to a 16% higher hazard (HR = 1.16, 95% CI: 1.04–1.29), and ≥2 disorders were associated with a 27% higher hazard (HR = 1.27, 95% CI: 1.06–1.52). Similarly, cardiometabolic burden was associated with increased CDW risk, with HRs of 1.20 (95% CI: 1.05–1.36) for one condition and 1.32 (95% CI: 1.07–1.65) for ≥2 conditions. When examined individually, all comorbidities except anxiety were significantly associated with a higher CDW risk (HR ranges: 1.12 for diabetes, 1.52 for peripheral vascular disease).

### Prediction of 3-month confirmed disability worsening (CDW) using MSM and Cox PH

We evaluated model performance in predicting 3-month CDW through internal validation with 500 bootstrap samples in each cohort. MSM-derived predictions of 3-month CDW showed closer alignment with the empirical Kaplan-Meier curve and lower IBS compared with Cox PH predictions in both cohorts (**Figure 4A-B**). The MSM achieved an IBS of 0.19 (95% CI: 0.16–0.22) vs. 0.27 (95% CI: 0.22–0.32) for Cox PH In PROMOTE, while the MSM achieved an IBS of 0.17 (95% CI: 0.15–0.19) vs. 0.24 (95% CI: 0.20–0.27) for Cox PH in CLIMB. These differences are expected given the structural advantages of MSMs for this task. Unlike Cox PH models, MSMs incorporate the full sequence of disability state transitions and intermediate states when generating CDW predictions, rather than modeling CDW as a single endpoint.

**Figure 4.**
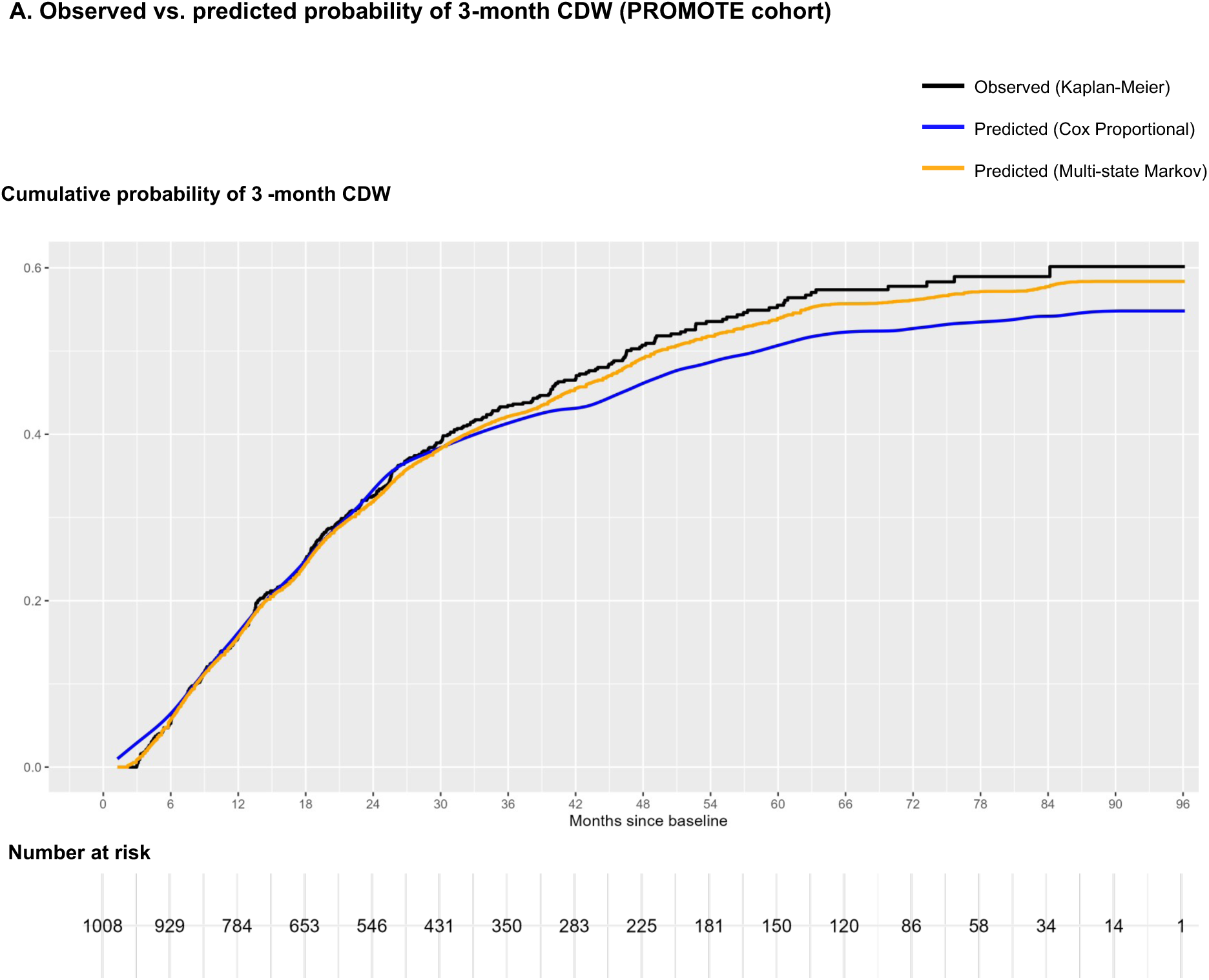
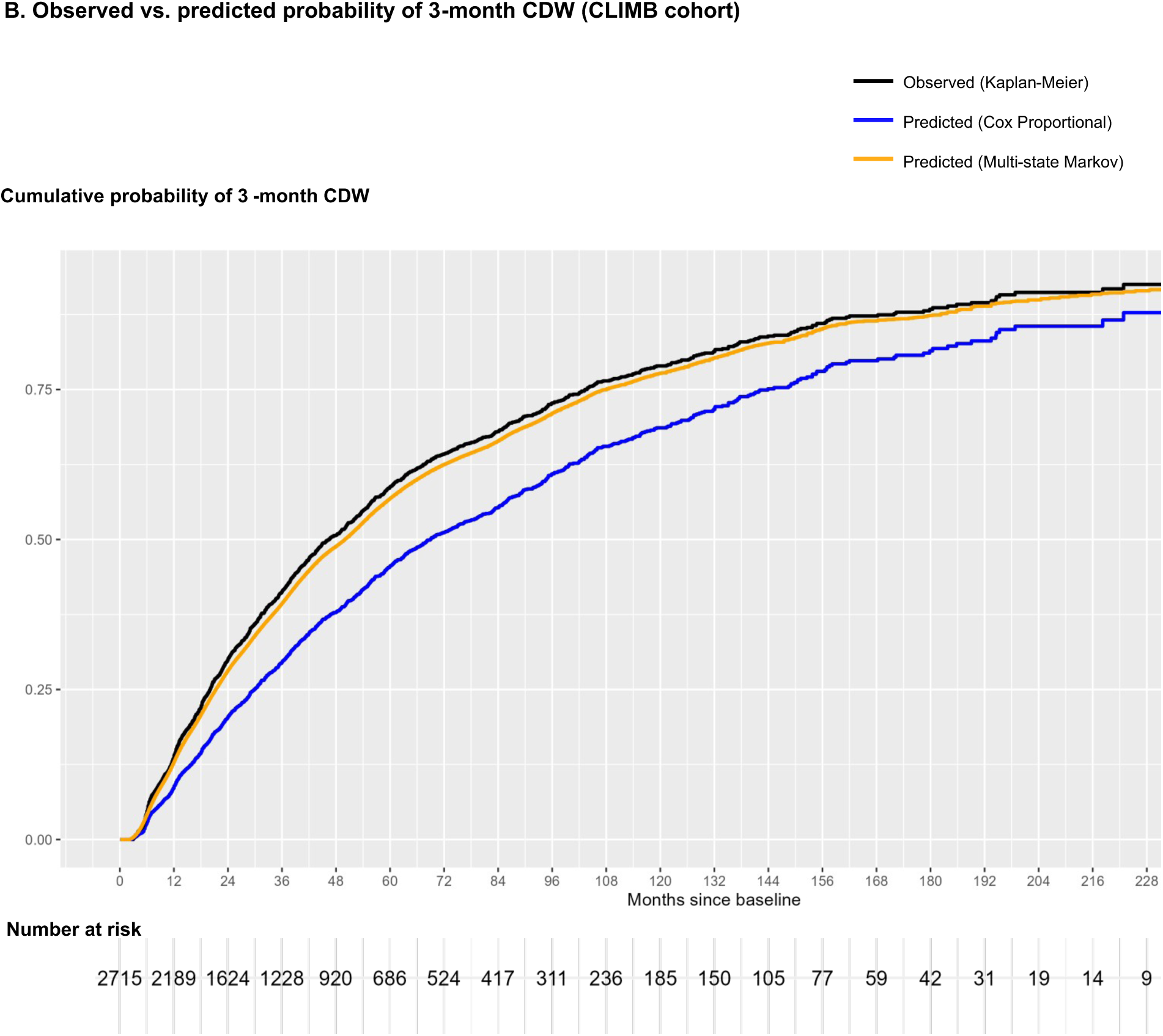
Predictions of 3-month confirmed disability worsening (CDW) in (A) PROMOTE cohort and (B) CLIMB cohort. The figure shows the overlay of Kaplan–Meier curve (black), Cox proportional hazard (Cox PH) model-based predictions (blue), and Multi-state Markov (MSM) model-based predictions (orange) of cumulative risk of 3-month CDW (y-axis) throughout the follow-up (x-axis). As observed, the MSM-based predictions more closely mirrored the Kaplan–Meier curve, indicating high prediction accuracy. Incorporating the same set of predictors, the mean Integrated Brier score (IBS) across bootstrap samples in the PROMOTE cohort was 0.19 for MSM and 0.27 for Cox PH models, and the mean IBS in the CLIMB cohort was 0.17 for MSM and 0.24 for Cox PH models.

### Sensitivity analyses

To evaluate the potential impact of missing comorbid diagnoses, we randomly assigned a proportion of participants without each condition to the corresponding comorbid condition group and found similar results (**eFigure 2A-D**). With a 5% random assignment, compared with patients without any comorbidities, patients with 1, 2, 3, and ≥4 comorbidities had 4%, 8%, 13% and 18% higher hazards of disability worsening (**eFigure 2A**), respectively, and 1%, 2%, 5% and 16% lower likelihood of disability improvement (**eFigure 2B**), respectively. Similar patterns were observed with a 20% random assignment (**eFigure 2C-D**).

## Discussion

Leveraging two large EHR-linked MS registries, this cohort study examined the associations of common comorbidities and overall comorbidity burden with disability transitions in 3,723 people with MS using MSMs. Unlike conventional survival models that focus on a single outcome event, MSMs captured clinically relevant, bidirectional disability transitions (i.e., both progression and improvement). We found that greater cardiometabolic and psychiatric comorbidity burden was associated with a higher likelihood of disability progression and a lower likelihood of disability improvement. Patients with ≥4 comorbidities had a 28% higher risk of worsening and a 20% lower likelihood of improvement than those without comorbidities. Each targeted comorbidity was significantly associated with worse disability outcomes, characterized by both higher worsening risk and lower improvement probability. These findings remained robust across sensitivity analyses accounting for potential comorbidity misclassification. Cox PH model results were directionally consistent, confirming that comorbidities were linked to worse outcomes when measured as CDW. However, Cox models showed inferior predictive performance for future CDW compared to MSMs.

To our knowledge, this is the first study to apply MSMs to quantify the comorbidity impact on disability dynamics in MS. Our findings align with prior clinical trial meta-analyses showing worse disability outcomes associated with greater comorbidity burden.^17^ By modeling both worsening and improvement simultaneously, we build on this literature and provide real-world evidence of how common cardiometabolic and psychiatric comorbidities shape MS disability trajectories.

MSM-derived predictions of CDW probabilities aligned more closely with observed outcomes than Cox PH-derived ones. This is not a critique of Cox PH modeling, which remains appropriate and well-suited for its intended purpose of estimating relative hazards for a single outcome event. Rather, when the clinical goal is to estimate absolute long-term CDW probability in dynamic outcome transition settings, a modeling framework that explicitly captures multi-state transitions yields more accurate predictions. This study supports the potential prognostic utility of MSMs in real-world MS settings, where disability evolves across multiple states over time.

This study examined common psychiatric and cardiometabolic comorbidities because of their high prevalence in MS, their designation as high priority by the International Advisory Committee on Clinical Trials in MS,^40^ and their clinical relevance as largely treatable conditions through multi-disciplinary care partnerships. Although phenome-wide screening can be useful for discovery,^14^ integrating such breadth with MSMs poses challenges, including complex multiple comparisons within the modeling strategy, limited power for low-prevalence conditions, and difficult temporal anchoring for non-chronic conditions. These challenges would amplify information and temporal biases. In contrast, our targeted strategy emphasizes clinically relevant, prevalent, and chronic conditions for which temporality is more reliably defined in EHRs, enabling a more rigorous assessment of their association with disability trajectories under a modeling framework that better reflects real-world dynamics.

In the two MS cohorts, anxiety, depression, hyperlipidemia, and hypertension were among the most common comorbid conditions. At baseline, about half of participants had no targeted comorbidities, similar to estimates from trial cohorts.^17^ However, the study cohorts had greater prevalence and higher total burden than trial cohorts (≥2 comorbidities: 28.4% vs. 18.2%), and higher domain-specific burdens (psychiatric ≥1: 32.7% vs. 17.4%; cardiometabolic ≥1: 30.6% vs. 23.3%). These differences suggest that trial populations often under-represent people with comorbidities due to stricter eligibility criteria and shorter follow-up durations, highlighting the value of real-world cohorts for quantifying comorbidity–disability relationships.

Modeling both worsening and improvement is essential because MS disability evolves along a continuum with phases of progression, remission, and recovery. Recent work using AI-driven reclassification of MS progression supports MS as a disease continuum.^18^ Our analysis complements this perspective by focusing specifically on disability states. Analyses examining solely worsening risk can underestimate the clinical importance of recovery, which can be influenced by biological repair processes and treatment effects. Our results indicate that the comorbidities studied not only accelerate progression but also hinder improvement, suggesting a dual pathway through which they shape disability trajectories.

Multiple mechanisms may plausibly explain our findings, including direct, indirect, and shared-cause pathways. For example, a Mendelian randomization study involving 12,584 MS cases reported a significant association between higher genetically determined body mass index (BMI) and increased MS disability, suggesting that obesity may directly exacerbate MS disability.^41^ Indirect pathways can also play a role, as comorbid conditions may interfere with disease management. A population-based Canadian study of 10,698 individuals with MS found that a higher total comorbidity burden (≥3 conditions vs. none) was associated with a lower likelihood of initiating higher-efficacy DMTs.^42^ Furthermore, the presence of any mental health disorder was associated with a 22% higher risk of earlier DMT discontinuation. Delayed initiation or discontinuation of DMTs may contribute to a higher risk of disability worsening and a lower chance of disability improvement. Shared environmental (e.g., air pollutants) and behavioral factors (e.g., smoking) increase the risk of both cardiometabolic conditions and MS disability accumulation.^43–45^ Finally, systemic inflammation underlying cardiometabolic and psychiatric conditions may exacerbate neurodegeneration and limit compensatory neuroplasticity, diminishing functional recovery. Together, these mechanisms provide a biologically and clinically coherent context for the observed dual effects.

Clinically, our findings have implications for prognosis. Existing disability prediction models in MS have primarily relied on extensive and multimodal inputs, including clinical assessments, laboratory biomarkers, and advanced neuroimaging metrics, most of which are not collected in routine clinical care, greatly limiting their practical adoption.^46,47^ Additionally, many models were trained on clinical trial cohorts and lose accuracy when applied to broader real-world populations.

While deep learning and other machine learning models have shown promise in improving prediction, they are often complex and lack interpretability. By contrast, MSMs retain transparency, are grounded in clinical measures (i.e., transitions between clinician-rated or patient-reported disability states), and, as shown here, outperform Cox models in predicting CDW. This positions MSMs as a promising, clinically interpretable alternative that could be integrated into routine prognostic tools, especially when comorbidity status can be readily ascertained from EHRs.

This study has several strengths. First, by employing MSMs, we modeled transition rates using the entire sequence of disability assessments, addressing limitations of conventional survival models, including their inability to capture bidirectional changes, susceptibility to competing risk bias, and reliance on a single event endpoint. Comparing results across modeling strategies, we found consistent evidence that comorbidities are associated with worse disability outcomes, underscoring the need for comorbidity management in MS. Moreover, MSMs demonstrated superior predictive performance for CDW, suggesting their potential as reliable clinical prognostic tools. Second, we ascertained comorbidities from objectively collected EHR data from two large healthcare systems, substantially reducing the recall bias that limited prior studies relying on self-reports. Third, we assessed the impact of potential comorbidity misclassification and showed that results remained robust. Finally, our analyses based on two large, independent EHR-linked clinic registries with demographically and clinically representative real-world MS populations (of the respective regions) extended the generalizability of findings regarding common comorbidities associated with MS disability transitions.

This study has limitations. First, the two study cohorts had heterogeneous findings, particularly in the associations of individual comorbidities with disability improvement. Between-cohort heterogeneity was generally low for worsening transitions but high (I²>50%) for several improvement transitions. This heterogeneity likely reflects differences in cohort characteristics: compared with CLIMB participants, PROMOTE patients were older, had longer disease duration, a higher prevalence of comorbidities, greater exposure to high-efficacy DMTs, shorter disability follow-up, and a higher likelihood of disability improvement. Although a random-effects model is generally preferred in meta-analyses with high heterogeneity, with only two cohorts we applied fixed-effects models to reduce unstable estimates of between-study variance. For several improvement transitions, the fixed-effects pooled HRs may not accurately represent a single underlying effect, and cohort-specific estimates are more informative.^48^ Future validation in additional MS populations is warranted to clarify the source and direction of this heterogeneity. Second, disability was measured differently between cohorts (PDDS vs. EDSS), and these scales are not isomorphic and do not correspond one-to-one. Our aim was *not* to directly compare results across cohorts but rather to harmonize scores into clinically meaningful disability states, enabling analysis of comorbidity effects on transitions between multiple states. Third, comorbidities were identified from EHR data using rule-based criteria (≥2 occurrences of corresponding PheCodes, separated by ≥3 days), which may introduce misclassification. Sensitivity analyses suggested that the observed results were robust to different levels of misclassification. Future work should explore machine learning-based phenotyping approaches using richer EHR data to more accurately capture comorbidity status and minimize information bias.^49^ Fourth, model performance was evaluated using internal validation via bootstrapping, which mitigated but may have not eliminated overfitting. External validation in independent cohorts will be necessary to confirm the superiority of MSMs over Cox PH models for predicting CDW. Finally, our MSM assumed nine disability states with only adjacent transitions allowed and employed a PR structure that constrains all progression transitions to share a common transition intensity. These restrictions ensured stable estimation with the available sample size and were consistent with prior MSM applications in MS research. However, with larger samples and longer follow-up — as both cohorts continue to accrue data — future work may relax these assumptions to better characterize how comorbidities shape disability dynamics across the full severity spectrum.

In summary, applying MSMs to two large EHR-linked clinic registry cohorts, we found that greater common cardiometabolic and psychiatric comorbidity burdens are associated with disability trajectories, predicting a higher risk of disability progression and a lower likelihood of improvement. By capturing both worsening and recovery transitions, MSMs provide clinically meaningful insights beyond conventional survival models and demonstrate superior predictive performance for CDW. These findings underscore the importance of routine comorbidity management in MS care and suggest the potential of MSMs as valuable prognostic tools in real-world settings.

## Supplementary materials

**eTable 1.** Individual comorbid conditions and associated PheCodes in this study.

| Category | Comorbidity | PheCode | PheCode String |
| --- | --- | --- | --- |
| Psychiatric | Depression | 296.2 | Depression |
|  |  | 296.22 | Major depressive disorder |
|  | Anxiety | 300 | Anxiety, phobic and dissociative disorders |
|  |  | 300.1 | Anxiety disorder |
|  |  | 300.11 | Generalized anxiety disorder |
|  |  | 300.2 | Generalized anxiety & phobic disorders |
|  |  | 300.3 | Obsessive-compulsive disorders |
|  | Other psychiatric disorders | 316 | Substance addiction and disorders |
|  |  | 296.1 | Bipolar |
|  |  | 301 | Personality disorders |
|  |  | 306 | Other mental disorder |
|  |  | 318 | Tobacco use disorder |
|  |  | 317 | Alcohol-related disorders |
| Cardiometabolic | Hypertension | 401 | Hypertension |
|  |  | 401.1 | Essential hypertension |
|  | Hyperlipidemia | 272.1 | Hyperlipidemia |
|  |  | 272.13 | Mixed hyperlipidemia |
|  | Cerebrovascular disease | 430 | Intracranial hemorrhage |
|  |  | 430.1 | Subarachnoid hemorrhage |
|  |  | 430.2 | Intracerebral hemorrhage |
|  |  | 430.3 | Subdural hemorrhage |
|  |  | 433 | Cerebrovascular disease |
|  |  | 433.1 | Occlusion and stenosis of precerebral arteries |
|  |  | 433.11 | Occlusion of cerebral arteries, with cerebral infarction |
|  |  | 433.12 | Cerebral atherosclerosis |
|  |  | 433.2 | Occlusion of cerebral arteries |
|  |  | 433.21 | Cerebral artery occlusion, with cerebral infarction |
|  |  | 433.3 | Cerebral ischemia |
|  |  | 433.31 | Transient cerebral ischemia |
|  |  | 433.32 | Moyamoya disease |
|  |  | 433.5 | Cerebral aneurysm |
|  |  | 433.6 | Acute, but ill-defined cerebrovascular disease |
|  | Functional cardiovascular disease | 411.2 | Myocardial infarction |
|  |  | 394.4 | Acute rheumatic heart disease |
|  |  | 394.2 | Mitral valve disease |
|  |  | 394.3 | Aortic valve disease |
|  | Ischemic heart disease | 411 | Ischemic heart disease |
|  |  | 411.1 | Unstable angina (intermediate coronary syndrome) |
|  |  | 411.3 | Angina pectoris |
|  |  | 411.4 | Coronary atherosclerosis |
|  |  | 411.8 | Other chronic ischemic heart disease, unspecified |
|  |  | 411.9 | Other acute and subacute forms of ischemic heart disease |
|  |  | 414 | Other forms of chronic heart disease |
|  | Peripheral vascular disease | 452.2 | Deep vein thrombosis |
|  |  | 456 | Chronic venous insufficiency |
|  |  | 443.8 | Other specified peripheral vascular diseases |
|  |  | 443.9 | Peripheral vascular disease, unspecified |
|  |  | 443 | Peripheral vascular disease |
|  | Diabetes | 249 | Secondary diabetes mellitus |
|  |  | 250 | Diabetes mellitus |
|  |  | 250.1 | Type 1 diabetes |
|  |  | 250.11 | Type 1 diabetes with ketoacidosis |
|  |  | 250.12 | Type 1 diabetes with renal manifestations |
|  |  | 250.13 | Type 1 diabetes with ophthalmic manifestations |
|  |  | 250.14 | Type 1 diabetes with neurological manifestations |
|  |  | 250.15 | Diabetes type 1 with peripheral circulatory disorders |
|  |  | 250.2 | Type 2 diabetes |
|  |  | 250.21 | Type 2 diabetes with ketoacidosis |
|  |  | 250.22 | Type 2 diabetes with renal manifestations |
|  |  | 250.23 | Type 2 diabetes with ophthalmic manifestations |
|  |  | 250.24 | Type 2 diabetes with neurological manifestations |
|  |  | 250.25 | Diabetes type 2 with peripheral circulatory disorders |

**eTable 2.** Categorization of disability states.

| Disability state | EDSS score | PDDS score |
| --- | --- | --- |
| State 0 | 0 | 0 |
| State 1 | 1 | 1 |
| State 2 | 1.5-2 | 2 |
| State 3 | 2.5-3 | 3 |
| State 4 | 3.5-4 | 4 |
| State 5 | 4.5-5 | 5 |
| State 6 | 5.5-6 | 6 |
| State 7 | 6.5-7 | 7 |
| State 8 | 7.5-8 * | 8 |

\* An EDSS score of 8.5–10 was not applicable, as no eligible participants had an entry state within that range. Note that EDSS and PDDS scores are not isomorphic and do not have a one-to-one correspondence. Instead, they were converted to clinically meaningful disability states, enabling analysis of comorbidity effects on transitions between states.

**eTable 3.** Variables and definitions used in the multi-state Markov models (MSM)

| **Variables** | **Definition** | **Time components** |
| --- | --- | --- |
| Individual comorbidity status<sup>1</sup> | Indicator for the presence of the specific comorbidity at the time of disability assessment (1 = Yes, 0 = No). | Time-varying. Once a participant met the criteria for comorbidity, they were considered to have that condition at the time of all subsequent disability assessments. |
| Comorbidity burden<sup>1</sup> | Total comorbidity count at each disability assessment, categorized as 0, 1, 2, or ≥3. Cardiometabolic and psychiatric burdens were each categorized as 0, 1, or ≥2. | Time-varying. Once a participant met the criteria for a burden category, it applied for the remainder of all subsequent disability assessments. |
| Sex | Self-reported at baseline (1 = Male, 0 = Female). | Time-independent |
| Race/ethnicity | Self-reported at baseline (1 = Non-Hispanic White, 0 = Other). | Time-independent |
| Age | Age at baseline. | Time-independent |
| Disease duration | Years from symptom onset to baseline. | Time-independent |
| Diagnosis delay | Years from symptom onset to MS diagnosis. | Time-independent |
| Treatment delay | Years from symptom onset to the earlier of either DMT initiation or baseline date. | Time-independent |
| Health care utilization | Total count of ICD codes and clinical encounter notes. | Time-varying |
| High-efficacy DMT use | Use of high-efficacy DMT at the disability assessment (1 = Yes, 0 = No). | Time-varying |
| Standard-efficacy DMT use | Use of standard-efficacy DMT at the disability assessment (1 = Yes, 0 = No). | Time-varying |
| MS subtype | Phenotype at the disability assessment (1 = Progressive MS, 0 = Relapsing-remitting MS). | Time-varying |
| Relapses | Total number of relapses up to the disability assessment. | Time-varying |

<sup>1</sup> Each exposure was evaluated individually as the primary independent variable in separate MSM models (i.e., they were not included together in a single model).

**eTable 4.** Variables and definitions used in Cox proportional hazard models (Cox PH)

| **Variables** | **Definition** | **Time components** |
| --- | --- | --- |
| Individual comorbidity status<sup>1</sup> | Indicator for the presence of the specific comorbidity during follow-up (1 = Yes, 0 = No). | Time-varying. Once a participant met the criteria for a comorbidity, they were considered to have that condition for the remainder of follow-up. |
| Comorbidity burden<sup>1</sup> | Total comorbidity count during follow-up, categorized as 0, 1, 2, or $\geq 3$. Cardiometabolic and psychiatric burdens were each categorized as 0, 1, or $\geq 2$. | Time-varying. Once a participant met the criteria for a burden category, it applied for the remainder of follow-up. |
| Baseline disability state | Disability state at baseline. | Time-independent |
| Sex | Self-reported at baseline (1 = Male, 0 = Female). | Time-independent |
| Race/ethnicity | Self-reported at baseline (1 = Non-Hispanic White, 0 = Other). | Time-independent |
| Age | Age at baseline. | Time-independent |
| Disease duration | Years from symptom onset to baseline. | Time-independent |
| Diagnosis delay | Years from symptom onset to MS diagnosis. | Time-independent |
| Treatment delay | Years from symptom onset to the earlier of either DMT initiation or baseline date. | Time-independent |
| Health care utilization | Total count of ICD codes and clinical encounter notes. | Time-varying |
| High-efficacy DMT use<sup>2</sup> | Use of high-efficacy DMT during follow-up (1 = Yes, 0 = No). | Time-varying |
| Standard-efficacy DMT use<sup>2</sup> | Use of standard-efficacy DMT during follow-up (1 = Yes, 0 = No). | Time-varying |
| MS subtype<sup>2</sup> | Phenotype during follow-up (1 = Progressive MS, 0 = Relapsing-remitting MS). | Time-varying |
| Relapses<sup>2</sup> | Total number of relapses during follow-up. | Time-varying |

<sup>1</sup> Each exposure was evaluated individually as the primary independent variable in separate MSM models (i.e., they were not included together in a single model).
<sup>2</sup> Time-varying covariates were incorporated using a start–stop (interval) data structure, where each observation interval represented a period during which covariate values remained constant.

**eTable 5.**
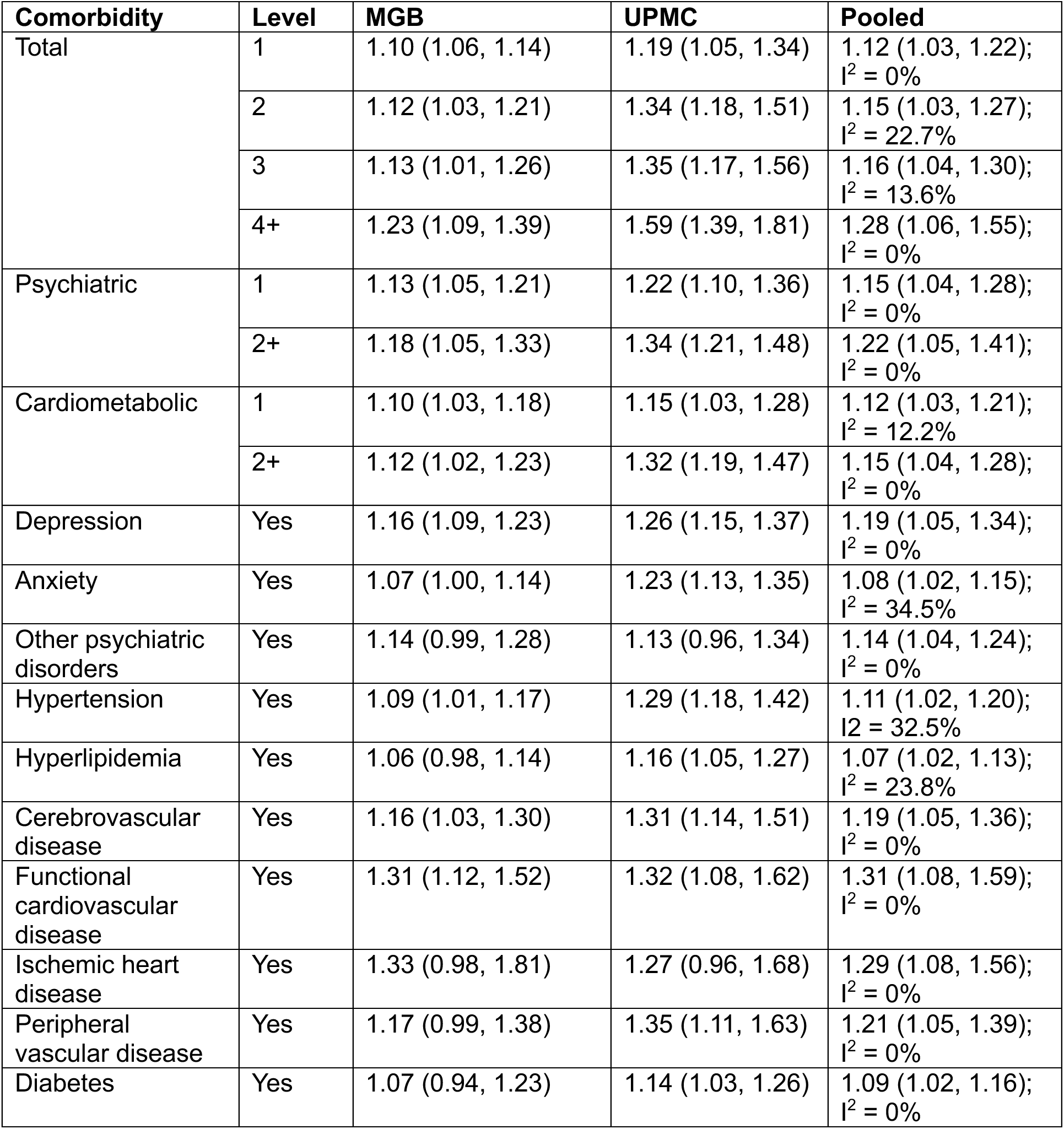
Hazard ratios and 95% confidence intervals for disability <u>worsening</u> by comorbidity status estimated from multi-state Markov models.

| **Comorbidity** | **Level** | **MGB** | **UPMC** | **Pooled** |
| --- | --- | --- | --- | --- |
| Total | 1 | 1.10 (1.06, 1.14) | 1.19 (1.05, 1.34) | 1.12 (1.03, 1.22);<br>$I^2 = 0\%$ |
| | 2 | 1.12 (1.03, 1.21) | 1.34 (1.18, 1.51) | 1.15 (1.03, 1.27);<br>$I^2 = 22.7\%$ |
| | 3 | 1.13 (1.01, 1.26) | 1.35 (1.17, 1.56) | 1.16 (1.04, 1.30);<br>$I^2 = 13.6\%$ |
| | 4+ | 1.23 (1.09, 1.39) | 1.59 (1.39, 1.81) | 1.28 (1.06, 1.55);<br>$I^2 = 0\%$ |
| Psychiatric | 1 | 1.13 (1.05, 1.21) | 1.22 (1.10, 1.36) | 1.15 (1.04, 1.28);<br>$I^2 = 0\%$ |
| | 2+ | 1.18 (1.05, 1.33) | 1.34 (1.21, 1.48) | 1.22 (1.05, 1.41);<br>$I^2 = 0\%$ |
| Cardiometabolic | 1 | 1.10 (1.03, 1.18) | 1.15 (1.03, 1.28) | 1.12 (1.03, 1.21);<br>$I^2 = 12.2\%$ |
| | 2+ | 1.12 (1.02, 1.23) | 1.32 (1.19, 1.47) | 1.15 (1.04, 1.28);<br>$I^2 = 0\%$ |
| Depression | Yes | 1.16 (1.09, 1.23) | 1.26 (1.15, 1.37) | 1.19 (1.05, 1.34);<br>$I^2 = 0\%$ |
| Anxiety | Yes | 1.07 (1.00, 1.14) | 1.23 (1.13, 1.35) | 1.08 (1.02, 1.15);<br>$I^2 = 34.5\%$ |
| Other psychiatric disorders | Yes | 1.14 (0.99, 1.28) | 1.13 (0.96, 1.34) | 1.14 (1.04, 1.24);<br>$I^2 = 0\%$ |
| Hypertension | Yes | 1.09 (1.01, 1.17) | 1.29 (1.18, 1.42) | 1.11 (1.02, 1.20);<br>$I^2 = 32.5\%$ |
| Hyperlipidemia | Yes | 1.06 (0.98, 1.14) | 1.16 (1.05, 1.27) | 1.07 (1.02, 1.13);<br>$I^2 = 23.8\%$ |
| Cerebrovascular disease | Yes | 1.16 (1.03, 1.30) | 1.31 (1.14, 1.51) | 1.19 (1.05, 1.36);<br>$I^2 = 0\%$ |
| Functional cardiovascular disease | Yes | 1.31 (1.12, 1.52) | 1.32 (1.08, 1.62) | 1.31 (1.08, 1.59);<br>$I^2 = 0\%$ |
| Ischemic heart disease | Yes | 1.33 (0.98, 1.81) | 1.27 (0.96, 1.68) | 1.29 (1.08, 1.56);<br>$I^2 = 0\%$ |
| Peripheral vascular disease | Yes | 1.17 (0.99, 1.38) | 1.35 (1.11, 1.63) | 1.21 (1.05, 1.39);<br>$I^2 = 0\%$ |
| Diabetes | Yes | 1.07 (0.94, 1.23) | 1.14 (1.03, 1.26) | 1.09 (1.02, 1.16);<br>$I^2 = 0\%$ |

**eTable 6.** Hazard ratios and 95% confidence intervals for disability <u>improvement</u> by comorbidity status estimated from multi-state Markov models.

| **Comorbidity** | **Level** | **MGB** | **UPMC** | **Pooled** |
| --- | --- | --- | --- | --- |
| Total | 1 | 0.86 (0.79, 0.93) | 0.99 (0.84, 1.17) | 0.99 (0.97, 1.01); $I^2 = 68.7\%$ |
| | 2 | 0.85 (0.77, 0.94) | 0.98 (0.82, 1.18) | 0.98 (0.95, 1.01); $I^2 = 66.1\%$ |
| | 3 | 0.76 (0.67, 0.87) | 0.96 (0.82, 1.12) | 0.95 (0.92, 0.99); $I^2 = 64.4\%$ |
| | 4+ | 0.56 (0.48, 0.66) | 0.83 (0.71, 0.97) | 0.8 (0.67, 0.95); $I^2 = 38.6\%$ |
| Psychiatric | 1 | 0.84 (0.76, 0.92) | 0.90 (0.78, 1.02) | 0.89 (0.82, 0.97); $I^2 = 0\%$ |
| | 2+ | 0.82 (0.75, 0.89) | 0.91 (0.85, 0.98) | 0.88 (0.80, 0.97); $I^2 = 0\%$ |
| Cardiometabolic | 1 | 0.81 (0.74, 0.89) | 0.99 (0.86, 1.14) | 0.99 (0.98, 1); $I^2 = 71.8\%$ |
| | 2+ | 0.65 (0.59, 0.73) | 0.88 (0.87, 0.99) | 0.87 (0.81, 0.94); $I^2 = 48.1\%$ |
| Depression | Yes | 0.82 (0.76, 0.89) | 1.00 (0.90, 1.12) | 0.94 (0.90, 0.98); $I^2 = 74.7\%$ |
| Anxiety | Yes | 0.89 (0.82, 0.96) | 0.89 (0.80, 0.99) | 0.89 (0.82, 0.97); $I^2 = 0\%$ |
| Other psychiatric disorders | Yes | 0.80 (0.69, 0.95) | 0.86 (0.71, 1.04) | 0.84 (0.74, 0.95); $I^2 = 0\%$ |
| Hypertension | Yes | 0.71 (0.65, 0.78) | 0.89 (0.80, 0.99) | 0.87 (0.78, 0.97); $I^2 = 34.3\%$ |
| Hyperlipidemia | Yes | 0.73 (0.66, 0.80) | 0.96 (0.86, 1.08) | 0.96 (0.92, 0.99); $I^2 = 64.1\%$ |
| Cerebrovascular disease | Yes | 0.77 (0.66, 0.89) | 0.97 (0.81, 1.17) | 0.97 (0.94, 0.99); $I^2 = 65.1\%$ |
| Functional cardiovascular disease | Yes | 0.60 (0.40, 0.90) | 0.90 (0.64, 1.26) | 0.88 (0.8, 0.98); $I^2 = 56.9\%$ |
| Ischemic heart disease | Yes | 0.69 (0.57, 0.84) | 0.99 (0.78, 1.26) | 0.99 (0.98, 1); $I^2 = 72.9\%$ |
| Peripheral vascular disease | Yes | 0.70 (0.56, 0.87) | 0.77 (0.61, 0.98) | 0.75 (0.6, 0.92); $I^2 = 0\%$ |
| Diabetes | Yes | 0.63 (0.53, 0.75) | 0.80 (0.67, 0.95) | 0.76 (0.62, 0.94); $I^2 = 0\%$ |

**eTable 7.** Predicted probability of transitioning into each state by comorbidity burden over 5 years.

| Comorbidity = 0 |  |  |  |  |  |  |
| --- | --- | --- | --- | --- | --- | --- |
|  | Year 0 | Year 1 | Year 2 | Year 3 | Year 4 | Year 5 |
| State 0 | 1.00 | 0.72 | 0.54 | 0.43 | 0.35 | 0.29 |
| State 1 | 0.00 | 0.13 | 0.18 | 0.19 | 0.19 | 0.18 |
| State 2 | 0.00 | 0.03 | 0.06 | 0.08 | 0.10 | 0.11 |
| State 3 | 0.00 | 0.02 | 0.04 | 0.06 | 0.07 | 0.08 |
| State 4 | 0.00 | 0.02 | 0.04 | 0.05 | 0.06 | 0.07 |
| State 5 | 0.00 | 0.02 | 0.04 | 0.05 | 0.06 | 0.07 |
| State 6 | 0.00 | 0.02 | 0.04 | 0.05 | 0.06 | 0.07 |
| State 7 | 0.00 | 0.02 | 0.04 | 0.05 | 0.06 | 0.07 |
| State 8 | 0.00 | 0.02 | 0.04 | 0.05 | 0.06 | 0.06 |
| Comorbidity = 1 |  |  |  |  |  |  |
|  | Year 0 | Year 1 | Year 2 | Year 3 | Year 4 | Year 5 |
| State 0 | 1.00 | 0.69 | 0.50 | 0.37 | 0.29 | 0.24 |
| State 1 | 0.00 | 0.14 | 0.19 | 0.19 | 0.19 | 0.17 |
| State 2 | 0.00 | 0.03 | 0.07 | 0.09 | 0.10 | 0.11 |
| State 3 | 0.00 | 0.02 | 0.04 | 0.06 | 0.07 | 0.09 |
| State 4 | 0.00 | 0.02 | 0.04 | 0.06 | 0.07 | 0.08 |
| State 5 | 0.00 | 0.02 | 0.04 | 0.05 | 0.07 | 0.07 |
| State 6 | 0.00 | 0.02 | 0.04 | 0.06 | 0.07 | 0.08 |
| State 7 | 0.00 | 0.02 | 0.04 | 0.06 | 0.07 | 0.08 |
| State 8 | 0.00 | 0.02 | 0.04 | 0.06 | 0.07 | 0.08 |
| Comorbidity = 2 |  |  |  |  |  |  |
|  | Year 0 | Year 1 | Year 2 | Year 3 | Year 4 | Year 5 |
| State 0 | 1.00 | 0.68 | 0.48 | 0.36 | 0.28 | 0.20 |
| State 1 | 0.00 | 0.13 | 0.18 | 0.19 | 0.19 | 0.16 |
| State 2 | 0.00 | 0.03 | 0.07 | 0.09 | 0.10 | 0.10 |
| State 3 | 0.00 | 0.04 | 0.04 | 0.06 | 0.07 | 0.10 |
| State 4 | 0.00 | 0.04 | 0.04 | 0.06 | 0.07 | 0.10 |
| State 5 | 0.00 | 0.03 | 0.05 | 0.05 | 0.06 | 0.10 |
| State 6 | 0.00 | 0.03 | 0.05 | 0.06 | 0.07 | 0.09 |
| State 7 | 0.00 | 0.02 | 0.05 | 0.06 | 0.07 | 0.09 |
| State 8 | 0.00 | 0.02 | 0.05 | 0.06 | 0.07 | 0.09 |
| Comorbidity = 3 |  |  |  |  |  |  |
|  | Year 0 | Year 1 | Year 2 | Year 3 | Year 4 | Year 5 |
| State 0 | 1.00 | 0.67 | 0.46 | 0.33 | 0.26 | 0.15 |
| State 1 | 0.00 | 0.11 | 0.15 | 0.17 | 0.18 | 0.12 |
| State 2 | 0.00 | 0.05 | 0.07 | 0.09 | 0.10 | 0.11 |
| State 3 | 0.00 | 0.03 | 0.05 | 0.08 | 0.10 | 0.09 |
| State 4 | 0.00 | 0.03 | 0.04 | 0.07 | 0.10 | 0.09 |
| State 5 | 0.00 | 0.03 | 0.05 | 0.07 | 0.09 | 0.10 |
| State 6 | 0.00 | 0.03 | 0.05 | 0.07 | 0.09 | 0.10 |
| State 7 | 0.00 | 0.03 | 0.05 | 0.06 | 0.09 | 0.11 |
| State 8 | 0.00 | 0.03 | 0.05 | 0.06 | 0.09 | 0.13 |
| Comorbidity ≥4 |  |  |  |  |  |  |
|  | Year 0 | Year 1 | Year 2 | Year 3 | Year 4 | Year 5 |
| State 0 | 1.00 | 0.66 | 0.45 | 0.32 | 0.24 | 0.15 |
| State 1 | 0.00 | 0.15 | 0.20 | 0.20 | 0.18 | 0.12 |
| State 2 | 0.00 | 0.04 | 0.07 | 0.10 | 0.11 | 0.11 |
| State 3 | 0.00 | 0.02 | 0.05 | 0.06 | 0.08 | 0.09 |
| State 4 | 0.00 | 0.02 | 0.04 | 0.06 | 0.07 | 0.09 |
| State 5 | 0.00 | 0.02 | 0.04 | 0.06 | 0.07 | 0.10 |
| State 6 | 0.00 | 0.02 | 0.05 | 0.06 | 0.08 | 0.10 |
| State 7 | 0.00 | 0.03 | 0.05 | 0.06 | 0.08 | 0.11 |
| State 8 | 0.00 | 0.03 | 0.05 | 0.08 | 0.10 | 0.13 |

**eTable 8.** Predicted total length of stay in each state by comorbidity burden during a 5-year period.

| | Comorbidity = 0 | Comorbidity = 1 | Comorbidity = 2 | Comorbidity = 3 | Comorbidity $\geq 4$ |
| --- | --- | --- | --- | --- | --- |
| State 0 | 3.05 | 2.46 | 2.42 | 2.23 | 2.03 |
| State 1 | 0.41 | 0.81 | 0.82 | 0.83 | 0.82 |
| State 2 | 0.32 | 0.35 | 0.35 | 0.38 | 0.40 |
| State 3 | 0.23 | 0.24 | 0.25 | 0.26 | 0.28 |
| State 4 | 0.21 | 0.22 | 0.23 | 0.24 | 0.26 |
| State 5 | 0.20 | 0.22 | 0.22 | 0.24 | 0.26 |
| State 6 | 0.20 | 0.22 | 0.23 | 0.25 | 0.28 |
| State 7 | 0.20 | 0.23 | 0.24 | 0.27 | 0.30 |
| State 8 | 0.19 | 0.24 | 0.25 | 0.31 | 0.37 |

**eTable 9.** Hazard ratios and 95% confidence intervals for <u>3-month confirmed disability</u> <u>worsening</u> by comorbidity status estimated from Cox proportional hazard models.

| **Comorbidity** | **Level** | **MGB** | **UPMC** | **Pooled** |
| --- | --- | --- | --- | --- |
| Total | 1 | 1.07 (0.85, 1.36) | 1.31 (1.00, 1.71) | 1.08 (0.99, 1.17); $I^2 = 50.9\%$ |
| | 2 | 1.17 (0.98, 1.40) | 1.59 (1.22, 2.07) | 1.21 (1.04, 1.4); $I^2 = 33.7\%$ |
| | 3 | 1.32 (1.15, 1.52) | 1.63 (1.19, 2.22) | 1.39 (1.09, 1.77); $I^2 = 0\%$ |
| | 4+ | 1.50 (1.14, 1.96) | 2.28 (1.71, 3.03) | 1.63 (1.13, 2.33); $I^2 = 0\%$ |
| Psychiatric | 1 | 1.13 (0.95, 1.35) | 1.27 (1.01, 1.60) | 1.16 (1.04, 1.29); $I^2 = 0\%$ |
| | 2+ | 1.22 (1.04, 1.42) | 1.57 (1.26, 1.97) | 1.27 (1.06, 1.52); $I^2 = 0\%$ |
| Cardiometabolic | 1 | 1.18 (1.01, 1.38) | 1.22 (0.95, 1.57) | 1.2 (1.05, 1.36); $I^2 = 0\%$ |
| | 2+ | 1.26 (1.05, 1.53) | 1.75 (1.40, 2.18) | 1.32 (1.07, 1.65); $I^2 = 11.4\%$ |
| Depression | Yes | 1.29 (1.12, 1.48) | 1.47 (1.21, 1.77) | 1.34 (1.09, 1.66); $I^2 = 0\%$ |
| Anxiety | Yes | 1.01 (0.87, 1.18) | 1.35 (1.12, 1.63) | 1.01 (0.98, 1.02); $I^2 = 71.9\%$ |
| Other psychiatric disorders | Yes | 1.28 (0.99, 1.64) | 1.30 (0.77, 1.87) | 1.29 (1.12, 1.5); $I^2 = 0\%$ |
| Hypertension | Yes | 1.20 (1.02, 1.40) | 1.57 (1.27, 1.91) | 1.25 (1.06, 1.47); $I^2 = 17.8\%$ |
| Hyperlipidemia | Yes | 1.18 (1.00, 1.38) | 1.31 (1.06, 1.62) | 1.21 (1.06, 1.39); $I^2 = 0\%$ |
| Cerebrovascular disease | Yes | 1.33 (1.03, 1.75) | 1.57 (1.13, 2.17) | 1.4 (1.09, 1.79); $I^2 = 0\%$ |
| Functional cardiovascular disease | Yes | 1.41 (0.70, 2.83) | 1.17 (0.62, 2.21) | 1.21 (1.05, 1.39); $I^2 = 0\%$ |
| Ischemic heart disease | Yes | 1.81 (1.25, 2.60) | 1.34 (0.83, 2.15) | 1.42 (1.10, 1.84); $I^2 = 0\%$ |
| Peripheral vascular disease | Yes | 1.42 (0.93, 2.15) | 2.29 (1.50, 3.49) | 1.52 (1.11, 2.10); $I^2 = 8.2\%$ |
| Diabetes | Yes | 1.10 (0.82, 1.48) | 1.45 (1.08, 1.95) | 1.12 (1.02, 1.23); $I^2 = 49.5\%$ |

**eFigure 1.**
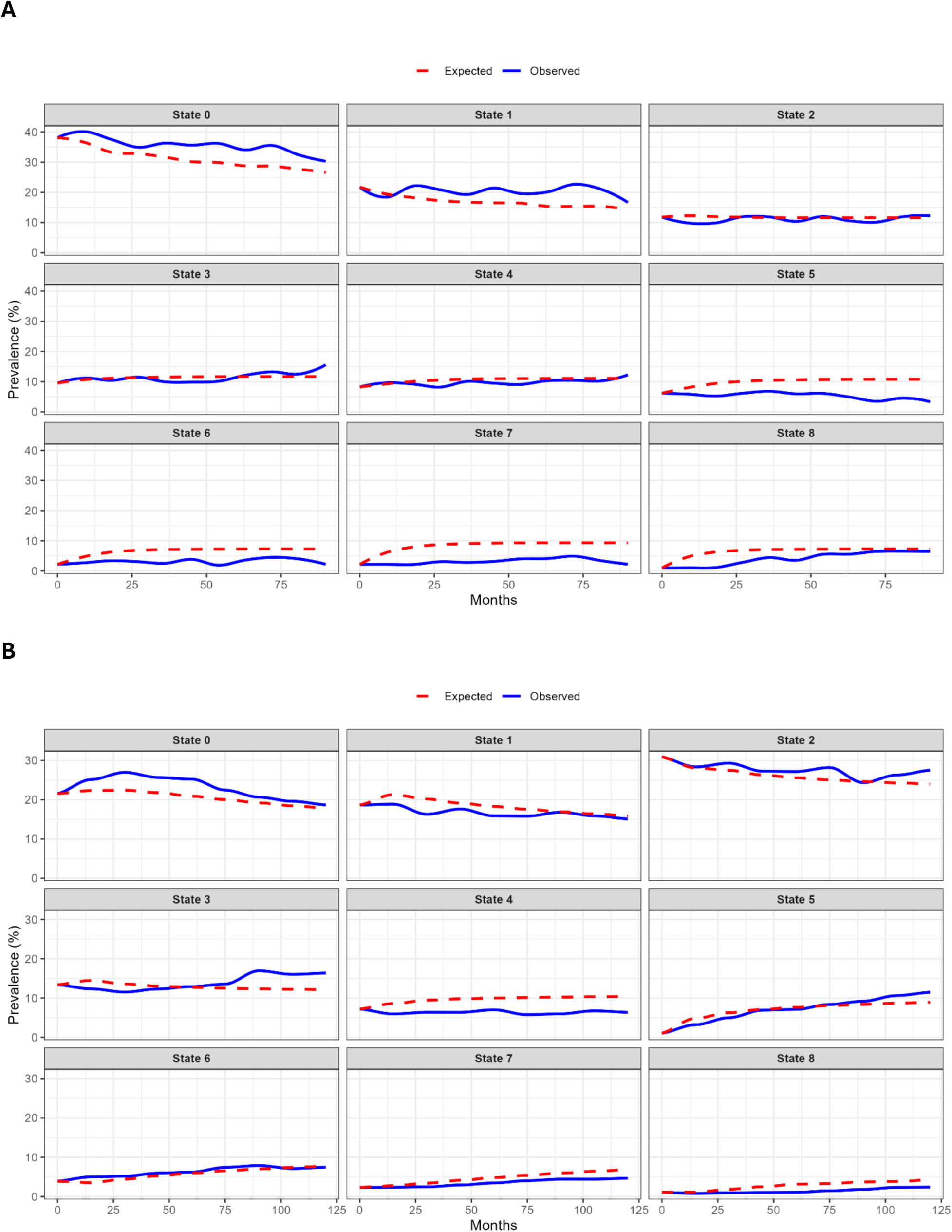
Observed and expected prevalences of patients occupying each state over time in (A) PROMOTE and (B) CLIMB cohort.

**eFigure 2.**
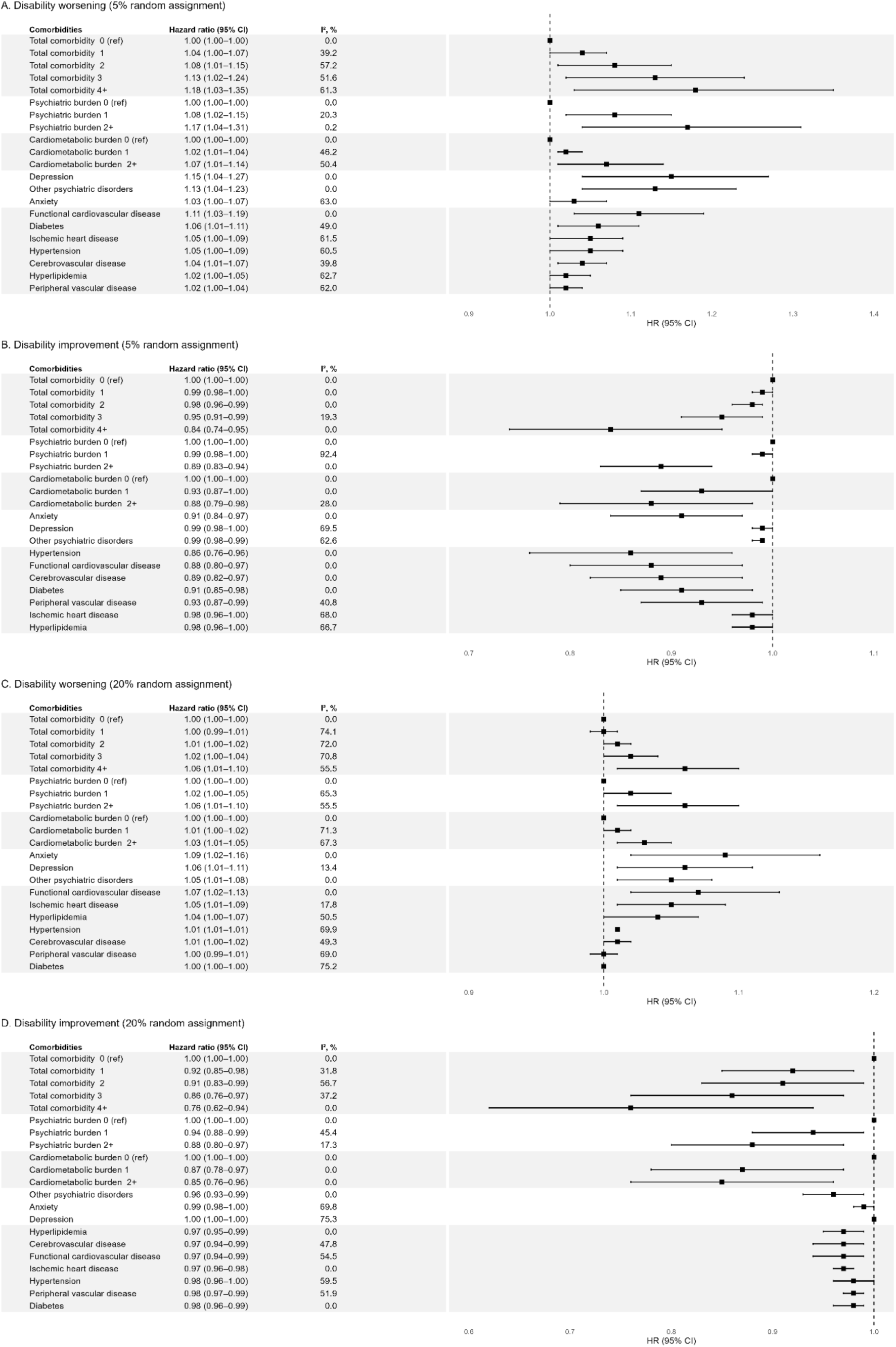
Pooled associations between comorbidities and disability outcomes: (A) Disability worsening with 5% random assignment (B) Disability improvement with 5% random assignment (C) Disability worsening with 20% random assignment, and (D) Disability improvement with 20% random assignment. All models adjusted for included comorbidities, sex, race/ethnicity, age, disease duration, diagnosis delay, treatment delay, healthcare utilization, DMT use, MS subtype, and total number of relapses.

## eMethods

### eMethod-1. DMT efficacy classification

Standard-efficacy DMTs included interferon beta-1a, interferon beta-1b, glatiramer acetate, fumarates (dimethyl fumarate, diroximel fumarate, monomethyl fumarate), daclizumab (before discontinuation), sphingosine-1-phosphate receptor modulators (fingolimod, siponimod, ozanimod, and ponesimod), and teriflunomide. Higher-efficacy DMTs included alemtuzumab, B-cell depletion agents (ocrelizumab, ofatumumab, rituximab, and ublituximab), cladribine, and natalizumab, as well as cyclophosphamide and mitoxantrone.

### eMethod-2. Penalized standard errors

To account for variability in estimation across multiple transitions with differing sample sizes, we applied a penalized standard error adjustment based on likelihood theory for second-generation p-values. Specifically, given a parameter estimate 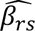 with standard error 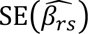 for the transition from state *r* to state *s*, we adjusted the standard error using the formula:

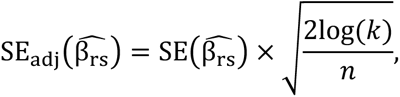

where *n* is the effective sample size for the transition (i.e., the number of observed transitions between states *r* and *s*), and *k* is the *likelihood support ratio* used to define the width of a likelihood support interval. For example, using a 1/8 support interval, a commonly used alternative to 95% confidence intervals, we set *k*=8, following Blume et al. (2019). This formulation reflects the principle that plausible parameter values lie within the region where the likelihood is no less 1/*k* of its maximum. The resulting intervals, known as likelihood support intervals, correspond to empirical Bayes 95% credible intervals under a non-informative prior.

This adjustment is not a classical multiple testing correction but rather a principled method for modulating inference precision based on data density and the relative evidential support for parameter values. As described in Blume et al. (2019), such likelihood-based intervals have superior frequency properties and align naturally with the framework of second-generation *p*-values, which aim to quantify whether the observed data support only null effects, scientifically meaningful alternatives, or are inconclusive. The penalty term log(*k*) governs the breadth of acceptable likelihood ratios, while dividing by *n* ensures wider intervals and greater uncertainty when data are sparse. This method improves robustness and reproducibility in multistate models by avoiding overly narrow confidence intervals and overstated certainty in poorly supported transitions.

### eMethod-3. Deriving 3-month confirmed disability worsening (CDW) probabilities from the Multi-State Model

To obtain the predicted probability of 3-month CDW from the MSM, we followed the procedure described by Mandel et al., with adaptations to our model specification. First, we estimated patient-specific transition intensities between disability states from the fitted MSM, using each person’s covariate values at time t (i.e., demographics, comorbidities, and clinical factors). The estimated coefficients and baseline transition intensities define a 3-month transition probability matrix P(x), where each entry *p*_*kj*_(*x*) represents the probability of moving from state k to state j over a 3-month interval, conditional on the current state and covariates x. Next, we constructed a modified (“working”) transition matrix to represent the CDW definition. We introduce an additional absorbing state 9 to represent CDW occurrence. For each starting disability state k, transition probabilities to states meeting the CDW criterion were reallocated to this absorbing state. All transitions into this absorbing state remained fixed in subsequent time steps: once entered, the process could not leave. For example, for k = 5, we defined a new state 9 and let p_69_ = p_66_ + p_67_ + p_68_, p_79_ = p_77_ + p_78_, p_89_ = p_88_, and p_66_ = p_67_ = p_68_ = p_77_ = p_78_ = p_88_ = 0. The entry in the 4th row and 7th column of this working matrix is therefore the probability of a CDW occurring 3 months or before. This approach leverages the Markov property of the MSM to use the full estimated transition process and ensures that predictions respect the operational definition of CDW in our study.

